# Connecting genomic and proteomic signatures of amyloid burden in the brain

**DOI:** 10.1101/2024.09.06.24313124

**Authors:** Raquel Puerta, Itziar de Rojas, Pablo García-González, Clàudia Olivé, Oscar Sotolongo-Grau, Ainhoa García-Sánchez, Fernando García-Gutiérrez, Laura Montrreal, Juan Pablo Tartari, Ángela Sanabria, Vanesa Pytel, Carmen Lage, Inés Quintela, Nuria Aguilera, Eloy Rodriguez-Rodriguez, Emilio Alarcón-Martín, Adelina Orellana, Pau Pastor, Jordi Pérez-Tur, Gerard Piñol-Ripoll, Adolfo López de Munian, Jose María García-Alberca, Jose Luís Royo, María Jesús Bullido, Victoria Álvarez, Luis Miguel Real, Arturo Corbatón Anchuelo, Dulcenombre Gómez-Garre, María Teresa Martínez Larrad, Emilio Franco-Macías, Pablo Mir, Miguel Medina, Raquel Sánchez-Valle, Oriol Dols- Icardo, María Eugenia Sáez, Ángel Carracedo, Lluís Tárraga, Montse Alegret, Sergi Valero, Marta Marquié, Mercè Boada, Pascual Sánchez Juan, Jose Enrique Cavazos, Alfredo Cabrera, Amanda Cano, Agustín Ruiz, Alzheimer’s Disease Neuroimaging Initiative

**Affiliations:** Ace Alzheimer Center Barcelona – Universitat Internacional de Catalunya, Spain; Universitat de Barcelona (UB); CIBERNED, Network Center for Biomedical Research in Neurodegenerative Diseases, National Institute of Health Carlos III, Madrid, Spain; Neurology Service, Marqués de Valdecilla University Hospital (University of Cantabria and IDIVAL), Santander, Spain; Grupo de Medicina Xenómica, Centro Nacional de Genotipado (CEGEN-PRB3-ISCIII). Universidade de Santiago de Compostela, Santiago de Compostela, Spain; Unit of Neurodegenerative diseases, Department of Neurology, University Hospital Germans Trias i Pujol, Badalona, Barcelona, Spain; The Germans Trias i Pujol Research Institute (IGTP), Badalona, Barcelona, Spain; Unitat de Genètica Molecular, Institut de Biomedicina de València-CSIC, Valencia, Spain; Unidad Mixta de Neurologia Genètica, Instituto de Investigación Sanitaria La Fe, Valencia, Spain; Unitat Trastorns Cognitius, Hospital Universitari Santa Maria de Lleida, Lleida, Spain; Institut de Recerca Biomedica de Lleida (IRBLLeida), Lleida, Spain; Department of Neurology. Hospital Universitario Donostia. San Sebastian, Spain; Department of Neurosciences. Faculty of Medicine and Nursery. University of the Basque Country, San Sebastián, Spain; Neurosciences Area. Instituto Biodonostia. San Sebastian, Spain; Alzheimer Research Center & Memory Clinic, Andalusian Institute for Neuroscience, Málaga, Spain; Departamento de Especialidades Quirúrgicas, Bioquímica e Inmunología. School of Medicine. University of Malaga. Málaga, Spain; Centro de Biología Molecular Severo Ochoa (UAM-CSIC); Instituto de Investigacion Sanitaria ‘Hospital la Paz’ (IdIPaz), Madrid, Spain; Universidad Autónoma de Madrid.; Laboratorio de Genética. Hospital Universitario Central de Asturias, Oviedo, Spain; Instituto de Investigación Sanitaria del Principado de Asturias (ISPA).; Unidad Clínica de Enfermedades Infecciosas y Microbiología.Hospital Universitario de Valme, Sevilla, Spain; Instituto de Investigación Sanitaria del Hospital Clínico San Carlos (IdISSC), Hospital Clínico San Carlos; Laboratorio de Riesgo Cardiovascular y Microbiota, Hospital Clínico San Carlos; Departamento de Fisiología, Facultad de Medicina, Universidad Complutense de Madrid (UCM).; Biomedical Research Networking Center in Cardiovascular Diseases (CIBERCV), Madrid, Spain; Centro de Investigación Biomédica en Red de Diabetes y Enfermedades Metabólicas Asociadas (CIBERDEM); Dementia Unit, Department of Neurology, Hospital Universitario Virgen del Rocío, Instituto de Biomedicina de Sevilla (IBiS), Sevilla, Spain; Unidad de Trastornos del Movimiento, Servicio de Neurología y Neurofisiología. Instituto de Biomedicina de Sevilla (IBiS), Hospital Universitario Virgen del Rocío/CSIC/Universidad de Sevilla, Seville, Spain; CIEN Foundation/Queen Sofia Foundation Alzheimer Center; Alzheimer’s disease and other cognitive disorders unit. Service of Neurology. Hospital Clínic of Barcelona. Institut d’Investigacions Biomèdiques August Pi i Sunyer, University of Barcelona, Barcelona, Spain; Department of Neurology, Sant Pau Memory Unit, Sant Pau Biomedical Research Institute, Hospital de la Santa Creu i Sant Pau, Universitat Autònoma de Barcelona, Barcelona, Spain; CAEBI, Centro Andaluz de Estudios Bioinformáticos, Sevilla, Spain; Fundación Pública Galega de Medicina Xenómica – CIBERER-IDIS, Santiago de Compostela, Spain; South Texas Medical Science Training Program, University of Texas Health San Antonio, San Antonio; Glenn Biggs Institute for Alzheimer’s & Neurodegenerative Diseases, University of Texas Health Science Center at San Antonio, 7703 Floyd Curl Dr, San Antonio, TX 78229 USA; Neuroscience Therapeutic Area, Janssen Research & Development, Turnhoutseweg 30, 2340 Beerse, Belgium

**Keywords:** Aβ42, CSF biomarkers, PET tomography, GWAS, Proteome

## Abstract

**Background:** Alzheimer’s disease (AD) has a high heritable component characteristic of complex diseases, yet many of the genetic risk factors remain unknown. We combined genome-wide association studies (GWAS) on amyloid endophenotypes measured in cerebrospinal fluid (CSF) and positron emission tomography (PET) as surrogates of amyloid pathology, which may be helpful to understand the underlying biology of the disease.

**Methods:** We performed a meta-analysis of GWAS of CSF Aβ42 and PET measures combining six independent cohorts (n=2,076). Due to the opposite effect direction of Aβ phenotypes in CSF and PET measures, only genetic signals in the opposite direction were considered for analysis (n=376,599). Polygenic risk scores (PRS) were calculated and evaluated for AD status and amyloid endophenotypes. We then searched the CSF proteome signature of brain amyloidosis using SOMAscan proteomic data (Ace cohort, n=1,008) and connected it with GWAS results of *loci* modulating amyloidosis. Finally, we compared our results with a large meta-analysis using publicly available datasets in CSF (n=13,409) and PET (n=13,116). This combined approach enabled the identification of overlapping genes and proteins associated with amyloid burden and the assessment of their biological significance using enrichment analyses.

**Results:** After filtering the meta-GWAS, we observed genome-wide significance in the rs429358-*APOE locus* and nine suggestive hits were annotated. We replicated the *APOE loci* using the large CSF-PET meta-GWAS and identified multiple AD-associated genes as well as the novel *GADL1* locus. Additionally, we found a significant association between the AD PRS and amyloid levels, whereas no significant association was found between any Aβ PRS with AD risk. CSF SOMAscan analysis identified 1,387 FDR-significant proteins associated with CSF Aβ42 levels. The overlap among GWAS *loci* and proteins associated with amyloid burden was very poor (n=35). The enrichment analysis of overlapping hits strongly suggested several signalling pathways connecting amyloidosis with the anchored component of the plasma membrane, synapse physiology and mental disorders that were replicated in the large CSF-PET meta-analysis.

**Conclusions:** The strategy of combining CSF and PET amyloid endophenotypes GWAS with CSF proteome analyses might be effective for identifying signals associated with the AD pathological process and elucidate causative molecular mechanisms behind the amyloid mobilization in AD.

## Background

Alzheimer’s disease (AD) is the most common cause of dementia. AD is a growing epidemic with an expected doubling of annual new diagnosis in the next 20 years prevalence and a major socioeconomic impact with a projected direct economic cost of $2 trillion by 2030^1–3^. In this sense, increasing the knowledge of AD aetiology and biomarker development would be an interesting approach to developing a clear understanding of the disease physiopathology and future drug developments. Genome wide association studies (GWAS) have permitted the discovery of more than 80 genetic variants associated with AD risk^4,5^. Despite the continued efforts led by international consortia, a large fraction of AD heritability remains to be elucidated since only 31% of AD genetic variance is explained by single-nucleotide polymorphisms (SNPs)^6^.

The analysis of heritable quantitative traits tightly linked to disease pathology, called endophenotypes, has become a promising approach in genetic studies^7–9^. These intermediate phenotypes might be influenced by the same genetic factors that confer risk to AD development and might have low genetic complexity. Compared to disease phenotypes, there are fewer genes or environmental influences affecting the genetic components of endophenotypes which facilitate finding a genuine association between these phenotypes and AD^7,8,10^.

The most common endophenotypes for AD are levels of amyloid beta (Aβ42), total tau (t-tau) and phosphorylated tau in threonine 181 (p-tau) in CSF^10–12^. Moreover, Positron Emission Tomography (PET) using several radiotracers for measuring amyloid and tau burden has been used as AD endophenotypes^11,13,14^. These biomarkers are surrogates of AD brain pathology and understanding their biology might provide insights into novel mechanisms of AD^15,16^. To date, relatively few AD *loci* have been identified using the endophenotype approach^9,13^. Moreover, GWAS analyses of PET and CSF endophenotypes are commonly analysed separately and comparisons between them have been overlooked.

In this study, we combined GWAS of Aβ CSF levels from four different AD cohorts with two GWAS of Aβ-burden measured using PET radiotracers. We used this strategy of combining both Aβ endophenotypes (CSF and PET) to identify novel genetic variants associated with AD and to replicate known AD signals. We then tested polygenic risk scores (PRS) derived from large studies in our datasets, dissected the CSF proteome signature associated with brain amyloidosis in a sizable CSF collection, and checked the overlapping of genomic meta-analyses and proteomic results.

## Materials and methods

### GWAS Cohorts

This study comprised a total of 2,076 individuals from Ace, Valdecilla and ADNI cohorts and had data for different Aβ CSF or PET endophenotypes (Supplementary Fig. 1 and Supplementary Table 1). To avoid overlap of subjects between the CSF and PET cohorts, we used only datasets with genotype-level information available.

#### a. Ace Alzheimer Center Barcelona

The Ace cohort comprised 1,189 individuals with brain amyloidosis measurements obtained using CSF or PET imaging, divided into three independent and non-overlapping cohorts. Because we used different methods to quantify CSF AB42, we decided to analyse the GWAS in two independent groups (536 individuals tested using Innotest ELISA kits and 472 individuals tested using the Lumipulse automated platform^17^). We included a third dataset of 181 individuals with subjective cognitive decline (SCD) tested using PET Florbetaben measures from the Fundació ACE Healthy Brain Initiative (FACEHBI) study^18^. The clinical protocols of the Ace Alzheimer Center have been previously published^17–19^. Briefly, syndromic diagnosis of all subjects was established by a multidisciplinary group of neurologists, neuropsychologists, and social workers. We assigned to healthy controls (HCs) including SCD diagnosis to Clinical Dementia Rating (CDR) of 0 individuals, and mild cognitive impairment (MCI) individuals a CDR of 0.5. For MCI diagnosis, the classification of López *et al.,* and Petersen’s criteria were also used^20–23^. We employed the 2011 National Institute on Aging and Alzheimer’s Association (NIA-AA) guidelines for AD diagnosis^24^. We performed a lumbar puncture (LP) to obtain CSF following consensus recommendations ^25^. The CSF obtained was centrifuged (2000 x g for 10 min at 4°C) and stored at -80°C. For Aβ42 analysis, CSF was defrosted at room temperature (20°C), vortexed and protein levels measured using the commercial ELISA kit Innotest β-AMYLOID (1-42) in 536 individuals and the chemiluminescent enzyme-immunoassay LUMIPULSE G600II automated platform (Fujirebio Europe, Belgium) in 472 individuals^17^. FACEHBI patients were assessed for brain amyloid deposition by PET imaging using the florbetaben [^18^F] radiotracer (FBB) (NeuraCeq©). A single slow intravenous bolus of 300 Mbq of FBB (6 sec/mL) (>10 mL during 20 min) was administered. After 90 min, PET images were acquired^18^.

#### b. Valdecilla cohort for the study of memory and brain aging

The Valdecilla cohort comprised 97 individuals who were older than 55 years and extensively phenotyped. Biological samples were collected at baseline and several tests were performed to evaluate early signs of AD. Moreover, core biomarkers in CSF were analysed and a neuropsychological battery including The Free and Cued Selective Reminding Test^26^, the Spanish version of the Face-Name Associative Memory Exam^27^, and the Logical Memory Test of the Wechsler Memory Scale-III^28^ and depression symptoms by the Geriatric Depression Scale^29^. HC (CDR=0), MCI (CDR=0.1) and dementia individuals (NIA-AA guidelines) were included in this analysis^30^. In the Valdecilla cohort, the Aβ42 biomarker was quantified by Lumipulse G600II which were interpreted according to previously established cut-off points^31^. Further information about phenotype assessment was presented elsewhere^32^.

#### c. Alzheimer’s Disease Neuroimaging Initiative (ADNI) cohort

Launched in 2003, ADNI is a longitudinal multicentre cohort for AD research based on United States and Canada^33,34^. The primary goal of ADNI has been to test whether biological markers, clinical and neuropsychological assessments can be combined to study the progression of MCI and early AD. We selected individuals from two separate ADNI databases: 1) the ADNI1 cohort with 378 individuals with available Aβ42 in CSF and 2) the ADNI2GO cohort with 412 individuals with available PET centiloid measures. In ADNI, syndromic diagnoses were based on a specific cut-off in the WMS-II LM test, education attainment, the Mini-Mental State Exam (MMSE) and CDR score. For HC and those with SCD, an MMSE score of 24–30 and a CDR of 0 were used. For those with MCI, a CDR of 0.5 and MMSE score of 24–30 were used. For those with AD, a CDR of 0.5–1 and an MMSE score of 20–26 were used. For the AD diagnosis, the National Institute of Neurological and Communicative Disorders and Stroke and Alzheimer’s Disease and Related Disorders Association (NINCDS/ADRDA) criteria for *probable* AD were considered^35^. In ADNI individuals, Aβ42 CSF biomarker was measured using the Luminex xMAP platform (Luminex Corp, Austin, TX) for multiplexing with Innogenetics immunoassay reagents (INNO-BIA AlzBio3, Ghent, Belgium)^36,37^. ADNI2GO patients were screened for brain amyloid deposits using the Florbetapir [^18^F] (AV45) radiotracer. After the injection of 370 MBq (10 mCi), four 5 min scans were acquired 50-70 min after the injection^36^. Further information about PET data acquisition can be found elsewhere^38^.

### PET imaging acquisition, harmonization and analysis

As FACEHBI and ADNI cohorts had different radiotracers, PET centiloid measures were used to perform a meta-analysis. Centiloids were calculated using equation (1), which was described for the conversion of FBB measures in the FACEHBI cohort^39^ and equation (2), which was described for the conversion of AV45 in ADNI^40^.

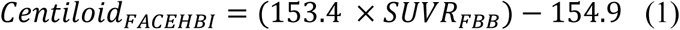

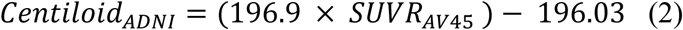

### Genotyping, quality control and imputation

Ace and Valdecilla DNA samples were genotyped using the Axiom 815K Spanish Biobank Array (Thermo Fisher). The genotyping was performed by the Spanish National Center for Genotyping (CeGen, Santiago de Compostela, Spain). Genotyping procedures have been previously published elsewhere^5,19^. For the ADNI samples, the Illumina Human610-Quad BeadChip platform was used for genotyping in ADNI1, and the Illumina HumanOmniExpress BeadChip was used for ADNI2GO^41^.

Common quality control was applied to all GWAS datasets. Briefly, individuals with low-quality samples, excess of heterozygosity, sample call rate below 97%, sex discrepancies, variants call rate below 95% or a deviation from the Hardy–Weinberg equilibrium (*P*>1e-06) were excluded from the analysis. In addition, familial relatedness (PI-HAT>0.1875) or ancestry outliers based on principal component analysis (PCA) were also removed. The imputation was performed using the Haplotype Reference Consortium panel in the Michigan Imputation Server^42^. Only the common markers (MAF>1%; MAC=20) and high imputation quality (R^2^>0.3) were selected for the subsequent analyses (GRCh37/hg19 reference assembly).

### SOMAscan Proteomic Assay

A subset of 1,008 CSF samples (Ace CSF cohort) was analysed using the SOMAscan panel measuring more than 7,000 proteins (SomaLogic, Boulder, Colorado). Briefly, this multiplex proteomic assay uses a 50 µL CSF sample and modified DNA aptamers to measure protein abundance. First, proteins are bound to immobilized aptamers using streptavidin beads and tagged with fluorescent markers. After washing unbounded proteins, the streptavidin beads are released using ultraviolet light, and the protein–aptamer complex is re-captured by monomeric avidin. To select only specific complexes, the protein–aptamers are exposed to an anionic competitor and then, hybridized in a conventional DNA array for analysis as described in Gold *et al*^43^. Finally, the protein level measures expressed in relative fluorescent units (RFU) are normalized using the adaptive normalization by maximum likelihood method further described in Candia *et al*^44^.

### Statistical analysis

#### a. GWAS and meta-GWAS

We harmonized CSF and PET endophenotypes measures performing a log10 transformation to adjust to a normal distribution, and Z-score values were determined using the *scale* R function (*center*=TRUE, *scale*=TRUE) (Supplementary Fig. 2). We used R software version 4.1.1.

The GWAS on each dataset was run using a generalized linear model in the software PLINK2^45^. The statistical model considered population microstratification (four PCs), sex, age, and dementia status for the association analysis. We then performed an inverse-variance weighted meta-analysis on each amyloid burden endophenotype separately, Aβ42 (n=1,483) in CSF and amyloid PET imaging (n=593).

Thereafter, both Aβ endophenotypes were further combined into a single meta-analysis (n=2,076) using the sample size weighted method in METAL software. This approach integrates p-values from different studies, weighting them by the sample size of each cohort, which provides a way to combine evidence across studies without relying on the effect size direction^46^. This is particularly useful when dealing with datasets where the effect sizes are not directly comparable or when different methods are used to measure the same biological outcome, as is the case with PET and CSF amyloid measurements.

We chose this meta-analysis of p-values approach because the effect directions and methods applied to measure amyloid burden differ between PET and CSF assessments. Specifically, in AD, the two measurements exhibit opposite biological directions: decreased levels of CSF Aβ42 are associated with increased amyloid plaque deposition in the brain, as observed through PET imaging. PET measures amyloid burden through radiotracer retention, while CSF measures it through soluble Aβ42 levels, which decrease as amyloid plaques accumulate in the brain. Thus, directly comparing effect sizes across these methods could be misleading^46,47^.

By combining p-values, focusing on the statistical significance and opposite effect size direction, this approach accounts for the differing biological contexts and measurement techniques, enabling a more robust and generalized analysis of amyloid burden across different datasets. The genetic markers evaluated in the meta-analysis were filtered considering the opposite effect direction in each CSF and PET endophenotype-independent GWAS and its presence in at least half of the datasets to select SNPs for further analysis.

Additionally, we performed another CSF-PET meta-analysis considering the largest publicly available datasets for CSF Aβ42 (n=13,116)^9^ and amyloid PET (n=13,409)^48^ (publicly available datasets; PAD analysis). To homogenize the results with our primary analysis, those datasets were converted to the GRCh37 assembly using the UCSC LiftOver software^49^. Because we did not have access to genotype-level information for all cohorts included in these studies, we were unable to prune potential overlapping subjects between both meta-analyses. Therefore, the results of combining these large meta-GWAS should be interpreted cautiously and are considered primarily for generating additional evidence about the pathways observed in our main analysis, where subject overlap was checked at the genotype level and removed to create two genuinely independent datasets (CSF and PET).

Finally, we attempted to replicate previously published genes for AD described in Bellenguez *et al*^4^, the significant signals associated with amyloid burden reported by the EADB consortium^9^ and significant markers associated with neuropathological features described in Beecham *et al*^50^ (Supplementary Table 2, Supplementary Table 3, Supplementary Table 4, and Supplementary Material 1).

#### b. Functional examination of identified sentinel SNPs and linked genomic regions

Clumping and annotation of suggestive signals (P<1e-05) were performed using the software PLINK1.9^51^. Additional annotations of biological function and gene-mapping were performed using meta-analysis summary statistics using the online tool Functional Mapping and Annotation of Genome-Wide Association Studies (FUMA)^52^. We set the threshold for independent significant SNPs at P<1e-05, R^2^<0.05, separated by over 250 kb, and we used the 1000G Phase3 reference panel to analyse suggestive signals in European population. For functional annotation, SNPs were matched to available databases such as ANNOVAR, Combined Annotation Dependent Depletion (CADD) scores, RegulomeDB and chromatin states based on a hidden Markov model from the Roadmap Epigenomics Project. Significant hits were mapped to genes according to 3 methods: 1) Physical distance with a maximum of 10 kb from nearby genes in the reference assembly, 2) expression quantitative trait *loci* (eQTL) associations assigned to SNP in blood, vascular, heart, brain tissues and embryonic stem cell derived cells, and 3) three-dimensional DNA interactions with SNPs and other gene regions where promoters were considered to be 250 bp upstream and 500 bp downstream of the transcription starting site for chromatin interaction. Moreover, a gene-based analysis was performed using MAGMA v1.08 that assigned exclusively protein-coding genes (Ensembl build 85) to the top SNPs found. Only 11,807 genes were mapped, and the gene-wide significance was defined at P=0.05/11,807=4.235e-06. We also conducted FUMA annotations in the amyloid burden meta-analysis considering the largest meta-GWAS for amyloid PET and CSF reported to date^9,48^.

#### c. Polygenic Risk Scores (PRS)

We computed the AD PRS described in Bellenguez *et al* that considered 83 *loci*. However, some SNPs were not imputed or had a low imputation quality (R^2^<0.3), and we decided to calculate the AD PRS including genetic variants found in all imputed datasets (n=76; Supplementary Table 5). For PRS calculation, we added the gene dosages of these SNPs weighting by their effect size (beta coefficients); the allele analysed was matched to the reported allele (A1) by Bellenguez *et al* ^4^. Because some control samples were included in the first stage of the AD GWAS, we considered the independent effects reported in the second stage for the PRS calculation. Additionally, due to the large effect on AD risk and its well-established association with most AD endophenotypes, the *APOE locus* was excluded from all these PRS. We then tested its association with AD case-control status, CSF Aβ42 and p-tau endophenotypes, and PET amyloid burden measurements. We considered as a covariate the age, sex, and disease status only in associations with biomarkers. These analyses were performed separately in each cohort except for Valdecilla which was excluded due to reduced sample size, while Ace PET cohort was excluded in the case-control analysis because all individuals were cognitively unimpaired. Additionally, we considered the fixed effect meta-analysis model considering the heterogeneity threshold (I^2^) of 75% as high^53^.

We also calculated another PRS for A) AD^4^ (n=76 SNPs; Supplementary Table 5), B) CSF Aβ42 levels (n=30 SNPs; Supplementary Table 6) considering the genetic variants with a P<1e-05 described in Jansen *et al* ^9^, and C) an amyloid burden PRS considering suggestive variants found in our meta-analysis (combining endophenotypes filtering according to the effect size direction; n=9 SNPs; Supplementary Table 7) in GR@ACE cohort individuals, including 8,110 cases and 9,640 controls the same way as described above. Further information about the cohort has been previously published^5,19^. For PRS computation, the effect (beta coeffcients) and standard errors were estimated using the equations described by Zhu *et al*^54^. Again, we associated these scaled PRS with case-control data in non-overlapping individuals to assess if Aβ genetic determinants are also related to disease risk.

#### d. Association between biomarker levels and SOMAscan proteomics

We assessed the association between SOMAscan 7k proteomic panel and CSF Aβ42 levels (n=1,008) in the Ace CSF cohort. Briefly, SOMAscan proteomic measures were log10 transformed, outliers were removed at ±3 standard deviations from the mean and standardized using the scale R function with centring and scaling. For further analysis, we selected 2,682 proteins based on correlations between: 1) two independent SOMAscan assay analysing the same samples, and 2) comparing aptamer- and antibody-based proteomic platforms^55,56^. To identify proteins associated to CSF Aβ42, a linear regression model was performed on scaled CSF Aβ42 levels and proteomic measures. We considered disease status, sex, age, and the CSF biomarker technique as covariates. Subsequently, the top 500 ranking of significant proteins associated with CSF Aβ42 (False discovery rate; FDR<1.864e-05) was analysed in the WEB-based GEne SeT AnaLysis Toolkit (WebGestalt)^57^ to perform an over-representation analysis (ORA) considering several functional databases and the whole genome as built-in reference gene list following the idea of investigating the complete genome (GWAS and gene-based analyses) (Supplementary Table 8). We also performed an enrichment analysis on the complete subset of valid SOMAscan proteins (n=2,682) to evaluate the impact of platform analyte preselection and quality control process on the results obtained.

To explore the biological significance of the GWAS results, we displayed the overlap between *loci* controlling amyloidosis and the proteins significantly associated with CSF Aβ42 in the Ace CSF cohort using Venn diagrams. The top 500 proteome and genome hits selected from the CSF Aβ42 meta-GWAS described in Jansen *et al*, the meta-analysis of CSF-PET endophenotypes filtered by opposite effect size direction, and the gene-based MAGMA analysis performed by FUMA were identified and annotated. The top rankings were reduced to 345, 339, 457, 361 and 465 *loci* for the meta-GWAS by Jansen *et al*, our current CSF-PET meta-analysis and its gene-based MAGMA analysis, the PAD CSF-PET meta-GWAS and its gene-based MAGMA analysis^9,48^, respectively. These reductions were due to the presence of SNPs that were not annotated and could not be matched to UniProt codes (Supplementary Table 9, Supplementary Table 10, Supplementary Table 11, Supplementary Table 12 and Supplementary Table 13). The top rankings were compared and the overlap between genomic and proteomic analysis was identified and evaluated using WebGestalt tool as described above.

## Results

### Meta-analysis of Aβ endophenotypes

The genome-wide meta-analysis of CSF endophenotypes involved 4 independent AD cohorts with Aβ42 measures (n=1,483; λ=1.009). The genomic inflation factor (λ) suggested no gross bias or stratification. As it was expected, we observed a consistent genome-wide significant association with rs429358-*APOE locus* as a sentinel variant (Effect=-0.58 [-0.658, -0.503]; P= 8.36e-49). We detected 19 additional suggestive pQTL signals for Aβ42 levels in CSF (Supplementary Table 14). Similarly, the meta-analysis of amyloid PET endophenotype (n=593; λ=1.013), revealed a genome-wide significant association in the same sentinel variant in the opposite direction (rs429358-*APOE locus*; Effect=0.684 [0.555, 0.813]; P= 2.00e-25). An additional novel hit at rs72737013 close to the *ANXA1* gene (Effect=0.813 [0.528, 1.099]; P*=*2.39e-08) was detected. This gene is related to anti-inflammatory reactions, innate immune response, and inflammatory processes^58^, psychiatric disorders, brain volume^59,60^, and the degradation of Aβ species^61^. Additionally, there were 43 additional independent suggestive signals annotated for amyloid burden measured using PET (Supplementary Table 15).

We combined the summary statistics from both CSF and PET Aβ meta-analyses without considering the effect direction (n=2,076). Again, we confirmed the sentinel variant rs429358 to be the most significant *locus* in the *APOE* region. Other genetic variants emerged as GWAS-significant in this new meta-analysis. However, none of them were inversely associated with CSF and PET endophenotypes in all studies except for the rs429358-*APOE* marker. We considered these hits as false positive signals (Supplementary Table 16).

In looking for new suggestive signals beyond *APOE*, we extracted the subset of 376,599 SNPs with consistent opposite effect in CSF and PET analyses. After the SNP selection in the combined Aβ meta-analysis, the rs429358-*APOE* variant (P*=*9.50e-67) remained as the only GWAS-significant hit (Fig. 1A, upper) but nine additional suggestive consistent variants were identified in genes such as *NPY5R, TIAM2* or *MAGI2,* among others (Table 1). Additionally, the combination of Aβ endophenotypes enhanced the significant replication of several genetic markers previously described for AD^4^, CSF Aβ42 levels^9^ and neuropathological features^50^ (Supplementary Material 1, Supplementary Table 2, Supplementary Table 3, Supplementary Table 4).

**Figure 1.**
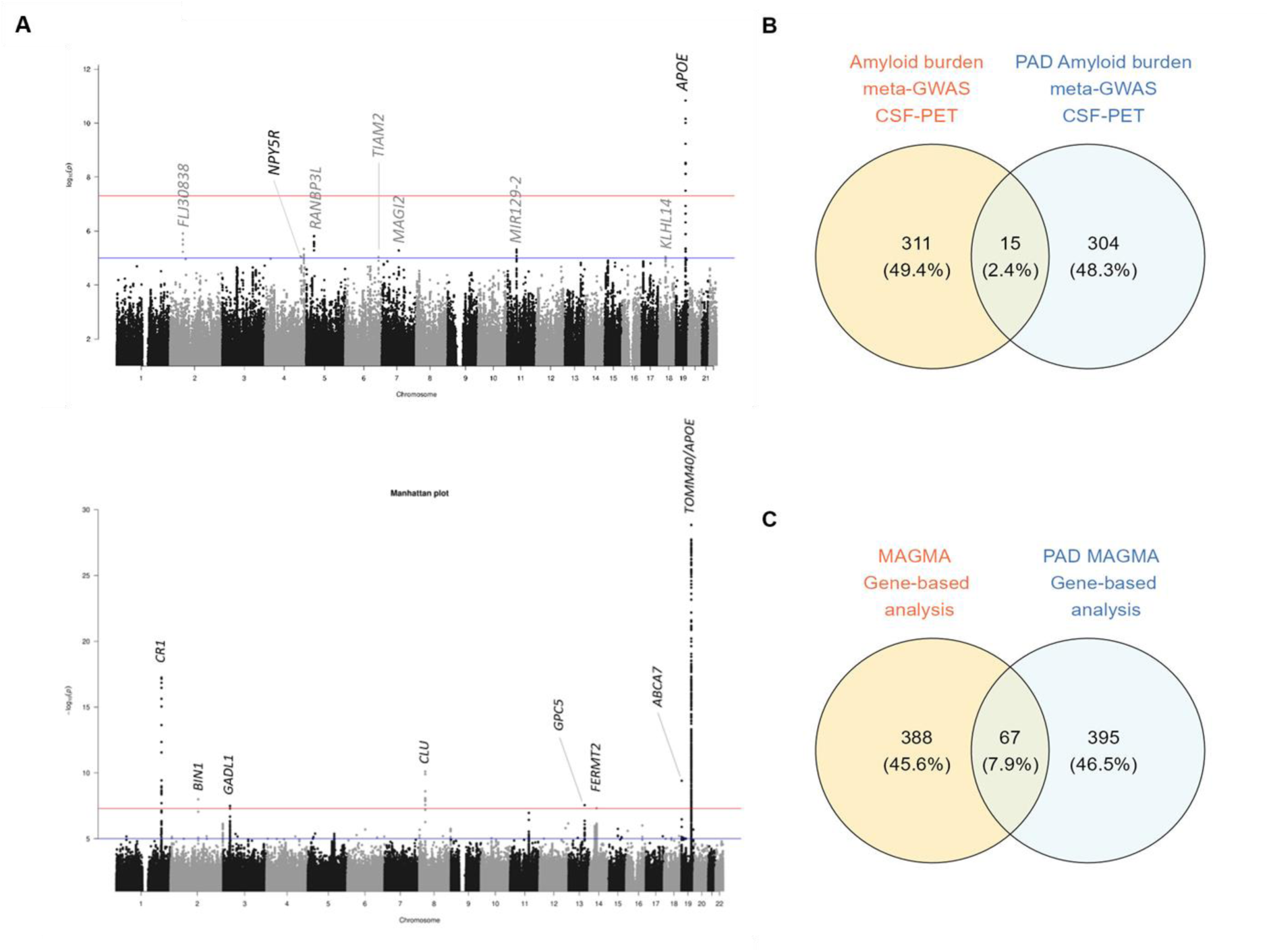
Plots of the Aβ burden meta-analysis combining data of CSF-PET endophenotypes. A) (upper) Manhattan plot of our CSF-PET meta-analysis (n=2,076). Results were filtered according effect size direction and dataset missingness. Suggestive independent markers were annotated with the nearest gene name. Mapped genes coloured in grey represent those that were not replicated in the PAD CSF-PET meta-GWAS. (lower) Manhattan plot of the PAD CSF-PET meta-analysis filtered (n=23,532). Genome-wide significant independent markers were annotated with the nearest gene name. The Y-axis was restricted to visualize suggestive signals. The genome-wide significance threshold was set to P<5e-08 (red line) and the suggestive threshold was set to P<1e-05 (blue line). B) Venn diagram representing the overlap between the top 500 ranking of independent genetic markers comparing the PAD and our amyloid burden meta-analysis. C) Venn diagram representing the overlap between the top 500 ranking of independent genes in the PAD and our gene-based analysis.

**Table 1.**
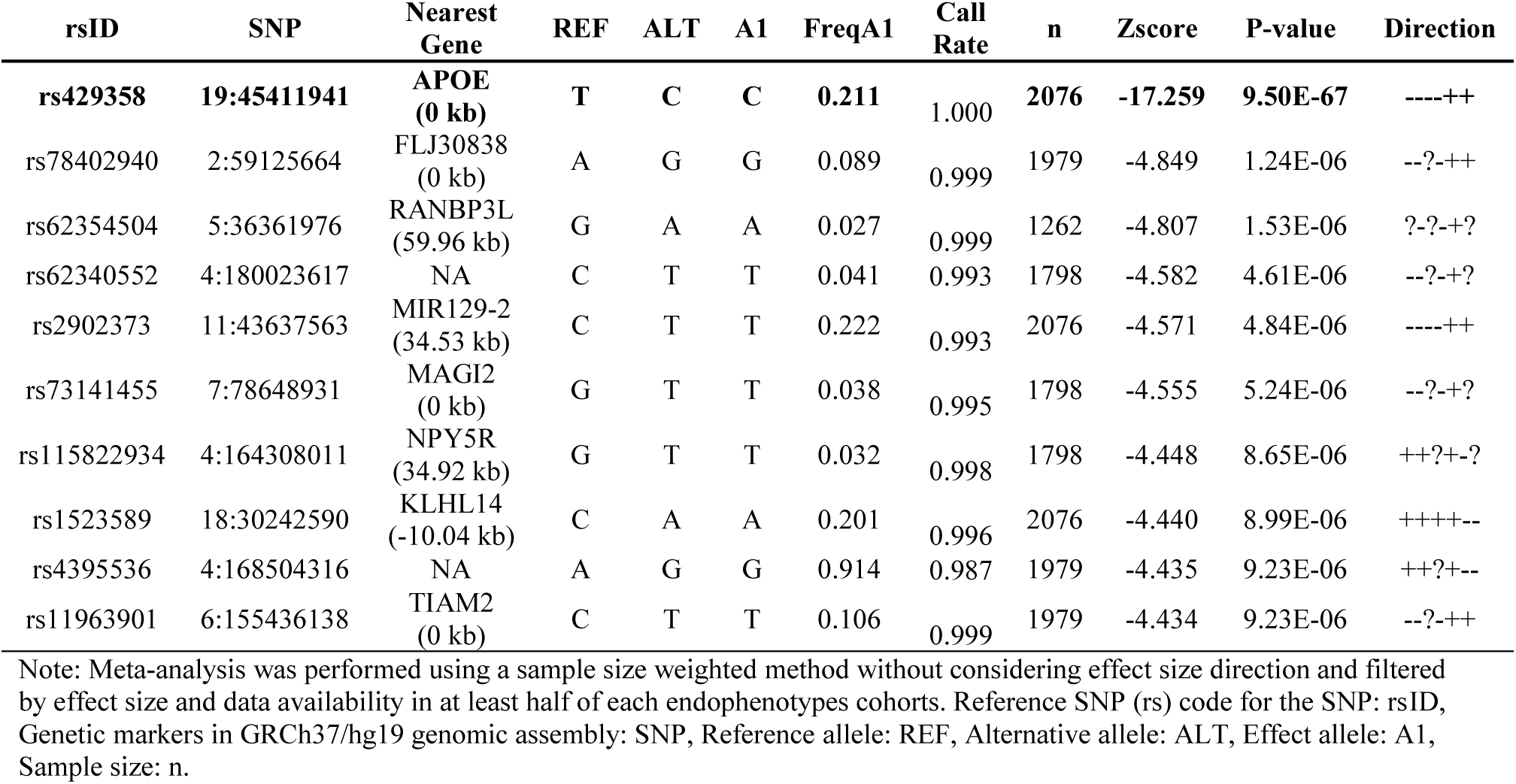
Results of the genome wide meta-analysis filtered combining CSF and PET endophenotypes (4.9% of total SNPs). Bold: significant results P < 5e-08 with consistent effect direction.

However, the PAD CSF-PET meta-analysis^9,48^ (effective sample size n=23,532) identified several markers previously associated with AD and its endophenotypes. These significant markers were identified on chromosome 19 including the rs429358-*APOE* (P=5.94e-601), as well as, the rs4844610-*CR1* (P=5.76e-18), rs7982-*CLU* (P=7.81e-11), rs12151021-*ABCA7* (P=3.92e-10), rs6733839-*BIN1* (P=1.02e-08), rs117834516-*FERMT2* (P=4.82e-08) and the novel rs4955351-*GADL1* (P=3.19e-08) which was not previously associated to AD or amyloid levels (Fig. 1A lower, Supplementary Table 17). Additionally, the PAD analysis replicated the rs115822934-*NPY5R* variant (P*=*3.21e-04) originally found suggestive in our CSF-PET meta-analysis (Supplementary Table 18). Importantly, we also observe concordances between our local effort (amyloid burden CSF-PET meta-GWAS) and the PAD. Specifically, we detected 15 overlapping sentinel markers in the top 500 ranking of the amyloid burden meta-GWAS from both the PAD and our current meta-analysis (Fig.1B), as well as 67 overlapping genes in the PAD and our gene-based top 500 ranking (Fig.1C).

To link the variants of interest to specific genes and obtain relevant functional information about these *loci*, we applied FUMA to the suggestive signals from the Aβ meta-analysis that were filtered based on opposite direction in CSF and PET (Table 1). There were 125 prioritized genes mapped using at least two strategies (positional mapping, eQTL or chromatin interactions) and 45 genes were selected based on the three strategies described in methods. As expected, the majority of the prioritized genes were related to the rs429358-*APOE*. Excluding chromosome 19, we prioritized 23 genes mapped (6 SNPs) with a CADD score > 12.37 suggesting a potential deleterious effect (Supplementary Table 19)^62^. In contrast to the univariate SNP analysis, the gene-based analysis performed using MAGMA revealed 15 study-wise significant *loci* (P<4.235e-06) excluding the *APOE* region (Table 2, Supplementary Fig. 3). Interestingly, the identified genetic variants in some of these genes (*TENM3*, *TMEM132D*, *PTPRD*, *CNTN5*, *RBFOX1*, *CSMD1*, *TIAM2*, *RORA* and *WWOX)* have been previously related to neuroimaging endophenotypes^63–65^, extreme AD PRS measures^66^, AD endophenotypes (CSF Aβ42 or p-tau levels^13,67–70^), mental disorders^71,72^ and cognitive decline in AD^64,73,74^. Additionally, the gene-based analysis of the PAD amyloid burden meta-GWAS revealed genes previously associated to AD such as *APOE* (P=2.09e-13), *CLU* (P=2.13e-07), *FERMT2* (P=3.49e-07) and the *CR1 locus* (P=3.64e-06), which reached borderline gene-wide significance threshold at P<2.717e-06 (Supplementary Table 20).

**Table 2.** Gene-based MAGMA results from FUMA analysis considering genome-wide significant results P<4.235e-06.

| Gene | EntrezID | UniProtID | CHR | Start | Stop | n<br>SNPS | Zscore | P-value |
| --- | --- | --- | --- | --- | --- | --- | --- | --- |
| NECTIN2 | 5819 | Q92692 | 19 | 45349432 | 45392485 | 37 | 7.787 | 3.442E-15 |
| APOE | 348 | P02649 | 19 | 45409011 | 45412650 | 4 | 7.282 | 1.648E-13 |
| APOC1 | 341 | P02654 | 19 | 45417504 | 45422606 | 5 | 6.774 | 6.278E-12 |
| CSMD1 | 64478 | Q96PZ7 | 8 | 2792875 | 4852494 | 929 | 6.437 | 6.076E-11 |
| TOMM40 | 10452 | O96008 | 19 | 45393826 | 45406946 | 24 | 6.109 | 5.000E-10 |
| WWOX | 51741 | Q9NZC7 | 16 | 78133310 | 79246564 | 323 | 5.566 | 1.307E-08 |
| LHPP | 64077 | Q9H008 | 10 | 126150403 | 126306457 | 153 | 5.507 | 1.822E-08 |
| BCL3 | 602 | P20749 | 19 | 45250962 | 45263301 | 6 | 5.449 | 2.540E-08 |
| CNTN5 | 53942 | O94779 | 11 | 98891683 | 100229616 | 556 | 5.358 | 4.217E-08 |
| TENM3 | 55714 | Q9P273 | 4 | 183065140 | 183724177 | 114 | 5.312 | 5.432E-08 |
| PTPRD | 5789 | P23468 | 9 | 8314246 | 10612723 | 332 | 5.258 | 7.272E-08 |
| TIAM2 | 26230 | Q8IVF5 | 6 | 155153831 | 155578857 | 60 | 5.243 | 7.904E-08 |
| RBFOX1 | 54715 | Q9NWB1 | 16 | 6069095 | 7763340 | 382 | 5.079 | 1.900E-07 |
| GPC5 | 2262 | P78333 | 13 | 92050929 | 93519490 | 275 | 4.956 | 3.604E-07 |
| RORA | 6095 | P35398 | 15 | 60780483 | 61521518 | 185 | 4.827 | 6.948E-07 |
| MACROD2 | 140733 | A1Z1Q3 | 20 | 13976015 | 16033842 | 354 | 4.787 | 8.471E-07 |
| TMEM132D | 121256 | Q14C87 | 12 | 129556270 | 130388211 | 205 | 4.759 | 9.717E-07 |
| CDH13 | 1012 | P55290 | 16 | 82660408 | 83830204 | 315 | 4.592 | 2.201E-06 |
| LRP1B | 53353 | Q9NZR2 | 2 | 140988992 | 142889270 | 318 | 4.549 | 2.696E-06 |
| KIR3DX1 | 90011 | Q9H7L2 | 19 | 55043909 | 55057053 | 9 | 4.487 | 3.616E-06 |
| FAM53B | 9679 | Q14153 | 10 | 126307861 | 126432838 | 42 | 4.479 | 3.751E-06 |

Chromosome: CHR, Number of SNPs: NSNP, Sample size: n, Entrez Gene Identifier: EntrezID, UniProt Swiss Protein Identifier: UniProtID.

### Association between AD PRS with AD endophenotypes and other clinical features

We observed a significant result in the meta-analysed associations between the AD PRS and Aβ levels (CSF Effect =-0.05 [-0.10, -0.00]; P*=*3.43e-02 and PET Effect =0.10 [0.02, 0.17]; P*=*1.30e-02). These results suggest that genes involved in AD risk indeed modulate amyloid levels (Fig. 2A, Fig. 2B).

**Figure 2.**
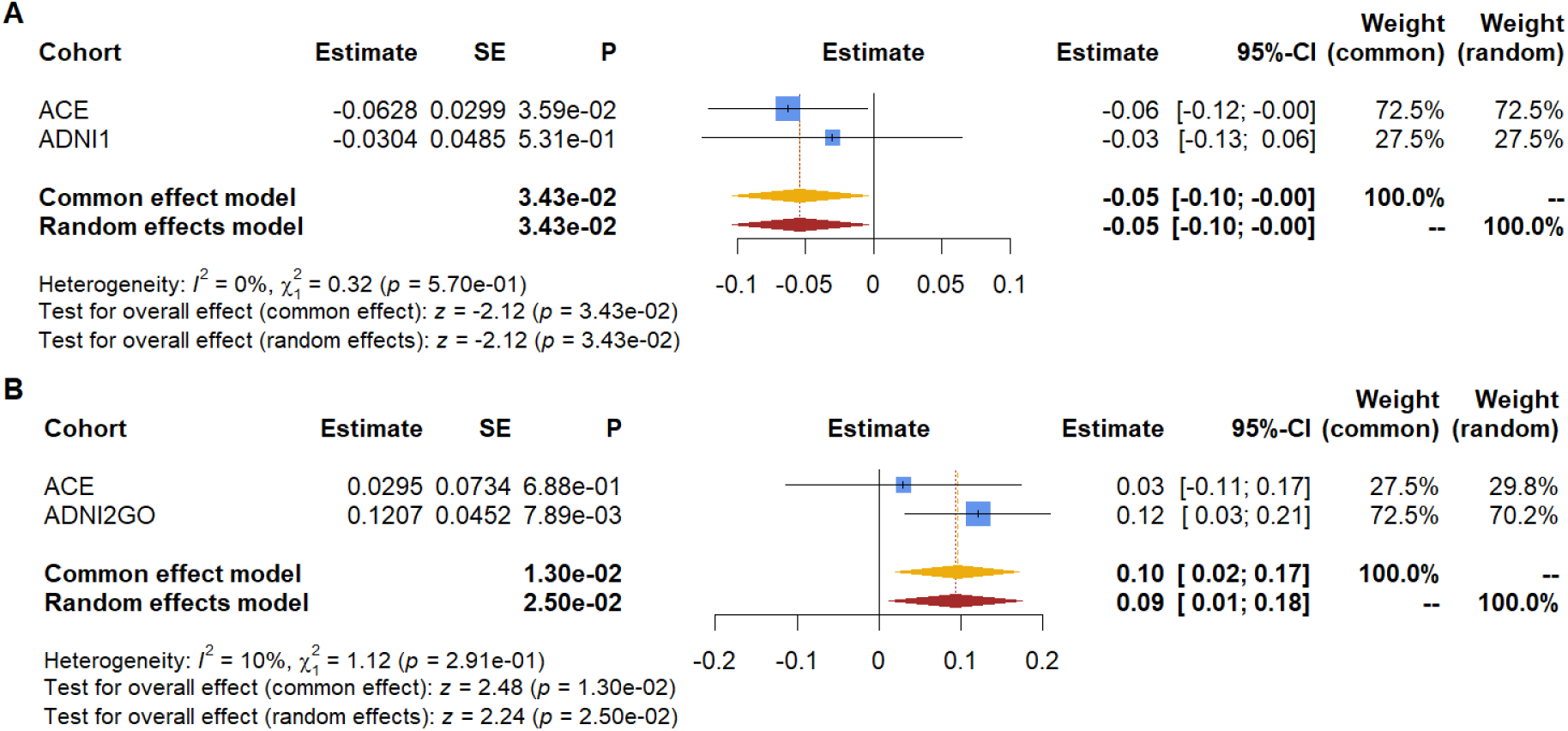
Forest plot of the meta-analysis association between the AD PRS. A) CSF Aβ42, and B) Aβ PET endophenotypes. The significance threshold was set to 0.05.

As expected, we observed a significant result in the association meta-analysis of the AD PRS with case-control dementia status in all 3 endophenotype datasets (OR=1.18 [1.05, 1.32]; *P=*5.29e-03). These results suggest that these genes modulate the disease status as previously reported^4^ (Fig. 3). Even though ADNI2GO did not reach statistical significance, it had a similar effect size and direction, possibly due to the low proportion of AD cases in this cohort (6.55 %).

**Figure 3.**
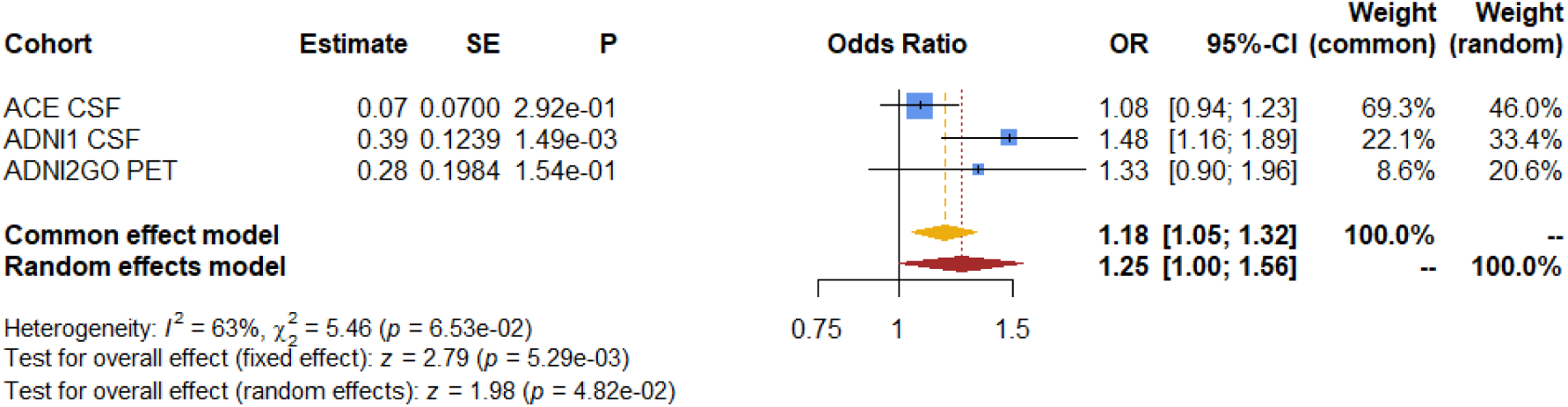
Forest plot of the meta-analysis association between the AD PRS and dementia status as case-control. In ACE (305 cases and 703 controls, 30.25%), ADNI1 (94 cases and 285 controls, 24.80%) and ADNI2GO cohorts (27 cases and 385 controls, 6.55%).

### Association between genetic variants of amyloid endophenotypes with case-control status

To assess whether CSF Aβ42 genetic modulators are also related to AD risk, we constructed different PRS including variants detected in our study and previous meta-GWAS^4^. We then checked the association of calculated PRS in the GR@ACE case-control study^5^. We did not observe any significant association for any calculated PRS for amyloid (Fig. 4) which could be due to the reduced set of independent markers reported for these phenotypes (P<1e-05) or not having an impact on AD pathology. As expected, the AD PRS was highly associated with the case-control (OR=1.35 [1.30, 1.40]; P*=*3.02e-49), thus confirming that the AD genes previously described by us and the EADB consortium^4,5^ truly modulate disease risk in the GR@ACE/DEGESCO cohort.

**Figure 4.**
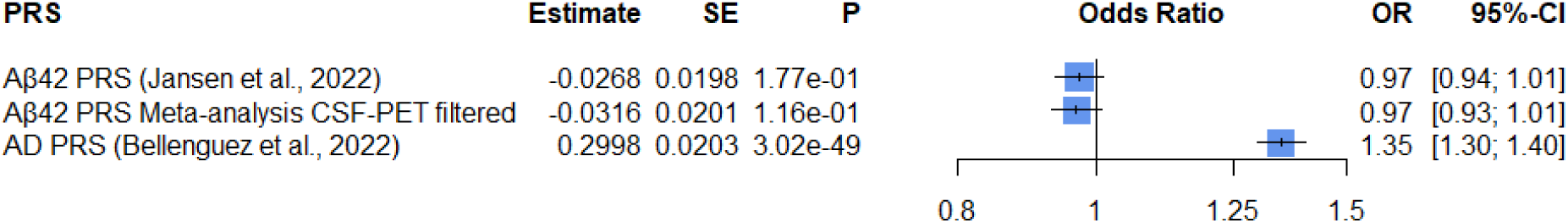
Forest plot of the association between the AD, Aβ PRS and case-control status. PRS for AD (76 SNPs from Bellenguez et al., 2022) and Aβ42 (30 SNPs from Jansen et al., 2022, 9 SNPs from our meta-analysis). The GR@ACE cohort included 8110 cases and 9640 controls.

### CSF proteome signatures associated with the Aβ42 CSF levels

We regressed the CSF Aβ42 peptide levels on CSF SOMAscan aptamer levels to identify the proteomic signature associated with amyloid burden (Fig. 5A). We identified 1,387 study-wide significant proteins in the linear model of CSF Aβ42 (FDR<1.864e-05) (Supplementary Table 21). Notably, we observed a marked asymmetry in the effect of SOMAmers on CSF Aβ42 levels, with the majority showing estimates greater than 0, suggesting a positive correlation contributing to increased CSF Aβ42 levels (Fig. 5A). Thus, the top 100 ranks of significant associations have an estimate range between 0.449 and 0.317, which contributes to an increase of this magnitude in CSF Aβ42 levels, and the variance explained by these highly associated proteins ranges of between 0.202 and 0.297. Importantly, we observed multiple proteins that have been associated with the CSF levels of Aβ species or its mechanisms in previous studies, such as MTMR7, LMOD4, GD3S, SERA/PHGDH, SELS, ATE1, NPTXR, and the 14-3-3 eta protein, among others^75–81^.

**Figure 5.**
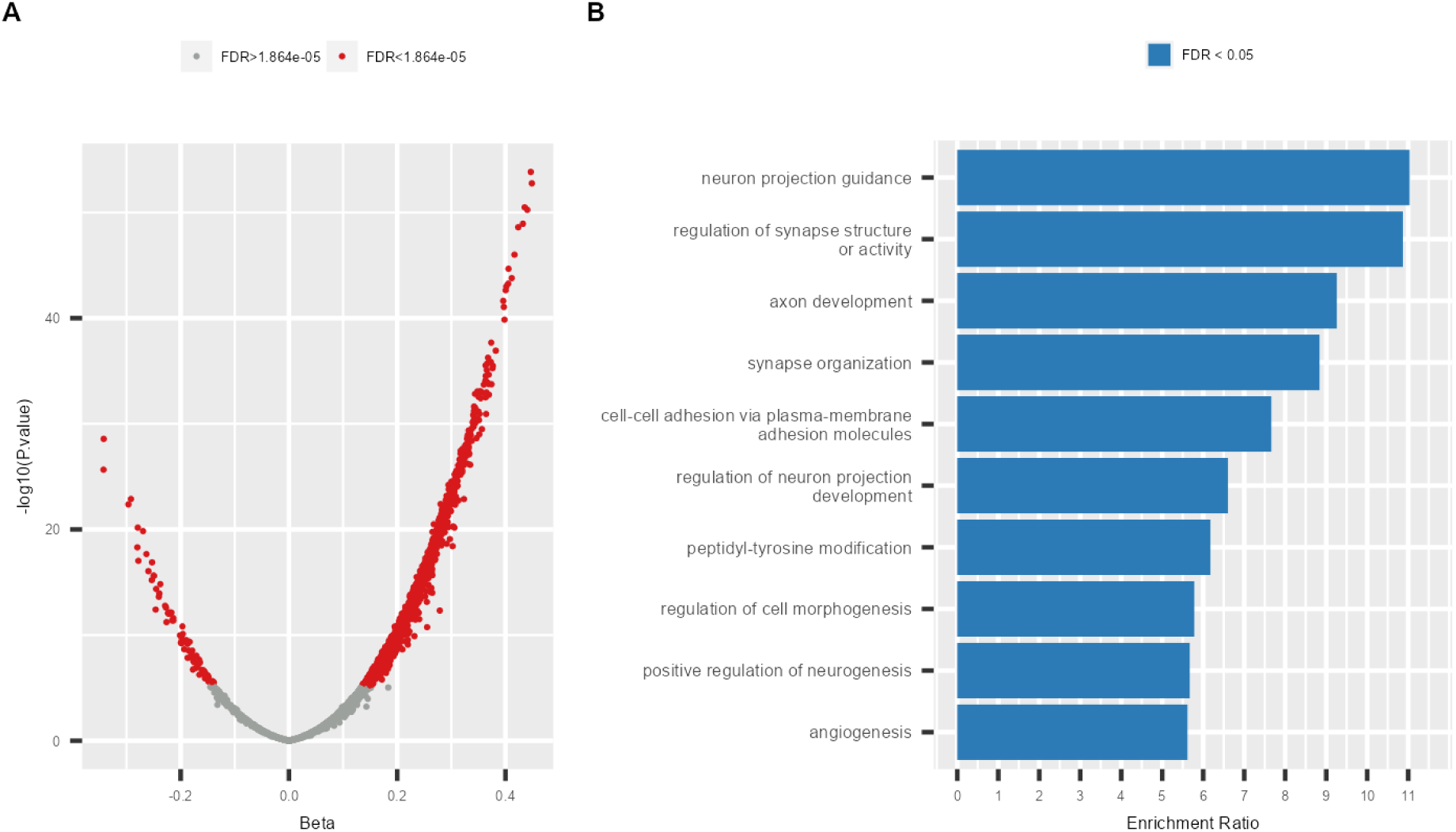
Associations between CSF SOMAscan and CSF Aβ42 levels. A) Vulcano plot only considering proteins with good inter-assay correlation (n=2,682), significant proteins (FDR < 1.864e-05) were highlighted in red (n=1,387). B) Top 10 results of the enrichment analysis of significant protein associations with CSF Aβ42 levels using the WebGestalt tool.

An enrichment analysis performed for significant proteins associated with CSF Aβ42 levels revealed genes involved in *neuronal projection guidance* (enrichment ratio=11.034; FDR<2.2e-16), *synaptic structure and activity* (enrichment ratio=10.868; FDR<2.2e-16), *cell– cell adhesion by plasma membrane molecules* (enrichment ratio=7.660; FDR<2.2e-16), *peptidyl-tyrosine modifications* (enrichment ratio=6.174; FDR<2.2e-16), *regulation of cell morphogenesis* (enrichment ratio=5.786; FDR<2.2e-16) and *angiogenesis* (enrichment ratio=5.617; FDR<2.2e-16) which are mainly driven by the large proportion of proteins with a positive effect (n=1,300; 93.73%) (Fig. 5B; Supplementary Table 22, Supplementary Table 23). Furthermore, when comparing the enrichment results from the ORA analysis between the entire set of valid SOMAscan proteins and the proteins significantly associated with Aβ42 levels, we observed a complete lack of overlap, reinforcing the validity of our findings (Supplementary Fig. 4).

To identify those genes that were commonly associated with CSF Aβ42 levels in genomic and proteomic analyses, we compared the top 500 common list of signals in the following four analyses: meta-GWAS by EADB^4^, our meta-analysis of CSF-PET, gene-based MAGMA, and SOMAscan protein analysis. We found three genes/proteins (*CHST1*, *PTPRD* and *TMEM132D*) present in all four analyses, representing only 0.2% of the total *loci*/proteins analysed (full overlap). In addition, 32 other proteins overlapped between the SOMAscan proteomics and any genomic analysis, including four proteins represented in 3 different analyses (Fig. 6A, Supplementary Table 24). Similar results were obtained in investigating the top rankings of the SOMAscan analysis, the PAD CSF-PET meta-GWAS and its gene-based analysis; only the *TMEM132D* was represented in all analyses (Fig. 6B). Interestingly, we found that 10 of the 23 *loci*/proteins observed were also overlapping with the ranking considering our main CSF-PET meta-GWAS results. This overlapping with PAD CSF-PET meta-GWAS support the validity of our approach (Supplementary Fig. 5, Supplementary Table 25). However, there was a reduced consistency between the top 500 SOMAscan proteins associated with CSF Aβ42 and any genomic results with less than 2.5% of overlapping proteins. These results suggest that the Aβ42-related protein signature in CSF might not be closely linked to amyloid genetic modulators, indicating that the proteome signature associated with Aβ42 burden in the brain primarily reflects general disease processes largely unrelated to the genetic elements controlling amyloid production.

**Figure 6.**
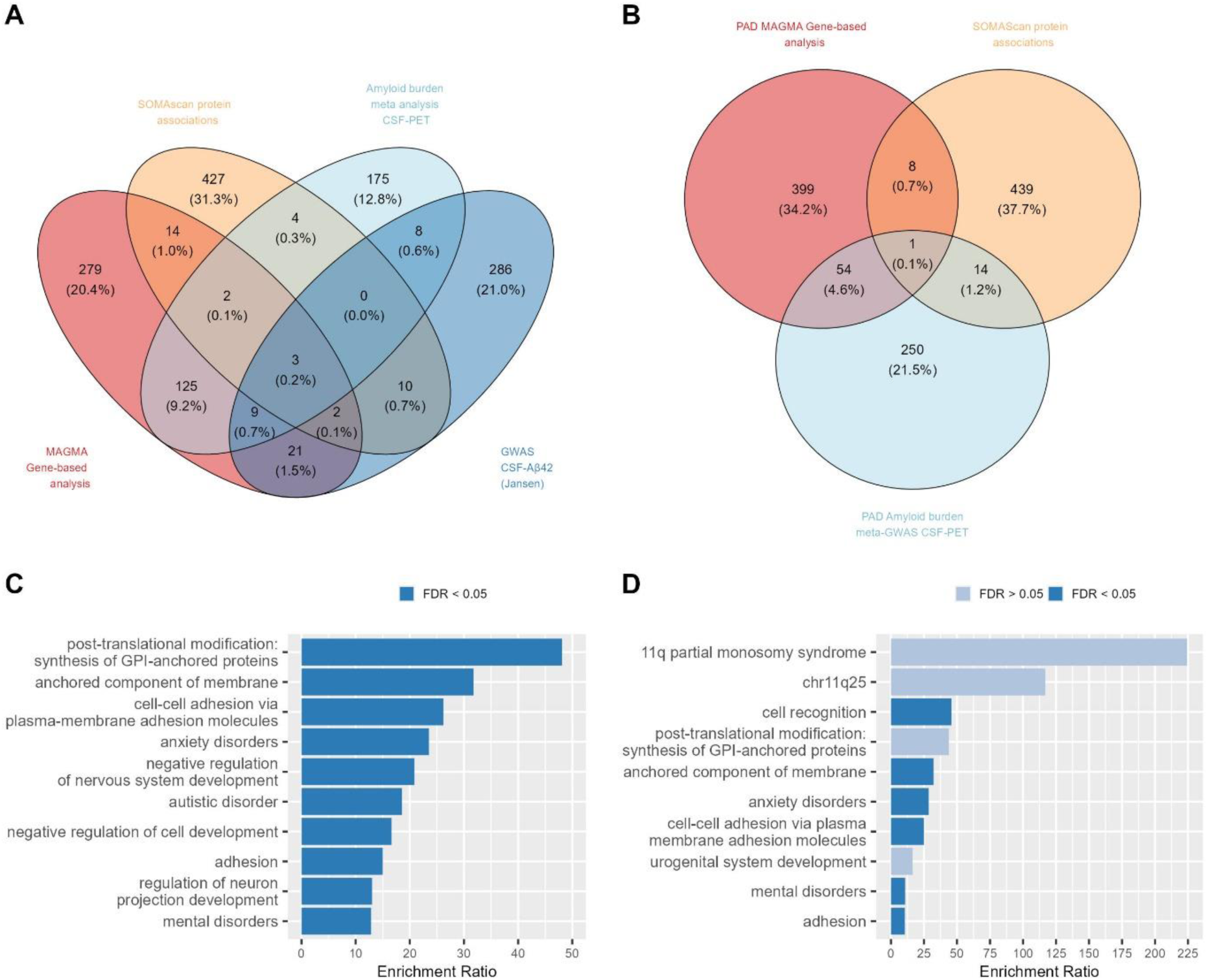
Overlapping loci/proteins in genomic and proteomic analysis. A) Venn diagram of the top 500 ranking of CSF Aβ42-associated proteins in the SOMAscan panel (orange), our gene-based MAGMA analysis (red), GWAS of CSF Aβ42 (Jansen et al., 2022) (dark blue) and our amyloid burden meta-analysis of filtered CSF-PET endophenotypes (light blue). B) Venn diagram of the top 500 ranking of CSF Aβ42-associated proteins in the SOMAscan panel (orange), PAD gene-based MAGMA meta-analysis (red) and PAD amyloid burden meta-analysis of filtered CSF-PET endophenotypes (light blue). C) Top 10 enrichment analysis results of the overlapping proteins between our genomic and proteomic analyses. C) Top 10 enrichment analysis results of the overlapping proteins between proteomic and PAD genomic analyses. The analysis was done using the WebGestalt tool.

Finally, to gain insight into the few commonalities identified by comparing genetic and proteome signatures associated to the amyloid burden in the brain, we conducted a new enrichment analysis. Despite the reduced overlapping hits among proteome and genome studies, several significant mechanisms related to the *synthesis of glycosylphosphatidyl inositol (GPI)-anchored proteins by post-translational modifications* were identified (enrichment ratio=48.070; FDR=1.86e-04) and the *anchored component of the membrane* (enrichment ratio=31.778; FDR=1.32e-04), *cell–cell adhesion via plasma membrane molecules* (enrichment ratio=26.207; FDR=3.42e-06), *mental disorders* (enrichment ratio=12.853; FDR=3.42e-06) such as *autism* (enrichment ratio=18.556; FDR=0.002) and *anxiety* (enrichment ratio=23.524; FDR=0.004), and *regulation and development of neuron projections* (enrichment ratio=13.035; FDR=0.002) among others (FDR<0.05). Interestingly, six of these mechanisms were also represented in the enrichment analysis of the PAD CSF-PET meta-analysis, which confirms our main results (Fig. 6C, Supplementary Table 26, Supplementary Table 27, Fig.6D, Supplementary Table 28, Supplementary Table 29).

## Discussion

For the first time, we have combined meta-GWAS results obtained from analysing amyloid PET and CSF Aβ42 levels. Our innovative experimental approach identified novel genetic variants associated with amyloid burden endophenotypes. This meta-analytic approach benefited from combining endophenotypic information from six cohorts thereby increasing our statistical power. As expected, we identified a genome-wide significant hit at the rs429358-*APOE loci*. We also observed a novel genome-wide significant hit near the *ANXA1 locus* exclusively associated with PET amyloid. SNPs in this *locus* were previously linked to psychiatric disorders, brain volume^59,60^, and the degradation of Aβ species^61^. However, neither the large PET meta-GWAS available nor the PAD meta-analysis conducted by us replicated this finding. For these reasons, we believe that this signal could be a false positive. We attribute the lack of additional hits to the relatively small sample size of our CSF-PET meta-GWAS. By repeating this strategy with a larger sample size, we expect to identify more genetic modulators of Aβ42 peptide expression in the brain. Indeed, using a similar approach with currently available summary statistics (PAD study), we were able to detect several sentinel markers surpassing the GWAS significance threshold. Specifically, the PAD CSF-PET meta-analysis identified several significant genes that have been previously related to AD (*CR1*, *BIN1*, *CLU*, *ABCA7*, *FERMT2* and *APOE*^4,82–85^) or amyloid proteins (*CR1*, *CLU*, *APOE* and *FERMT2*^9,86,87^), as well as *PICALM* and *GPC5* suggestive genes^84,88^. Notably, we identified the novel *GADL1* locus, which encodes for a protein from the glutamate decarboxylase family, suggesting that it might have a glutamate decarboxylase activity in the CNS^89,90^.

In AD, the glutamate excess generates a continuous glutamatergic activity, impairing neuronal plasticity and long-term potentiation leading to excitotoxicity. Therefore, using receptor antagonists such as memantine, which has shown neuroprotective effects, could be a crucial therapeutic intervention^91,92^.

Importantly, these results should be interpreted with extreme caution because PAD analysis is not entirely independent as various cohorts were represented in both summary statistics of the PAD analysis (Supplementary Table 30). This overlapping samples (11.284%) could lead to overestimated effects and increased proportion of false positive findings. Compared to our local effort, where we eliminated any potential overlap between CSF and PET cohorts, we remain very cautious about the PAD results due to the potential overlap of subjects among studies. Future efforts are necessary to confirm the findings from the PAD analysis. Nevertheless, the PAD analysis replicated the rs115822934-*NPY5R* marker, alongside the rs429358-*APOE*, originally identified in our CSF-PET meta-analysis. These results might suggest that *NPY5R* could be genuinely involved in amyloid pathology, as well as panic disorders^93,94^. Again, further completely independent studies, expanding the sample size of these analysis, are needed to validate our observation and working hypothesis.

In spite of these limitations, our experimental strategy permitted us to evaluate common pathways potentially associated to CSF-soluble Aβ42 (circulating amyloid)^95^ and brain amyloid species detected by PET (insoluble species such as amyloid plaques or cerebral amyloid angiopathy)^96^ and proteome signature associated to CSF Aβ42 peptide levels. To assess the relationship between genetic modulators and protein levels, we analysed the overlap between *loci*-controlling amyloid levels and significant proteins associated with CSF Aβ42 levels. Importantly, three genes/proteins (*CHST1*, *PTPRD* and *TMEM132D*) were identified and prioritized in all analyses, thus suggesting that these modulators might be key drivers controlling amyloid pathology. Lower *TMEM132D* levels have been observed in patients with frontotemporal dementia^97^, and genetic markers in this gene have been related to anxiety, panic disorders and the rate of cognitive decline^73,98,99^. This locus was the only that also overlapped with all PAD rankings, suggesting that might be a potential modulator of amyloid pathology. The *PTPRD* gene, which was also represented in the large meta-GWAS gene-based ranking, has been significantly associated to synaptic process in schizophrenia^100^, AD susceptibility, neurofibrillary tangle and neuritic plaques^68^. We consider these two *loci* excellent candidates for further translational research due to their consistent statistical significance and previous literature findings. Nevertheless, there is a possibility that we are not capturing pathological mechanisms occurring similarly in both biofluids due to the opposite direction filtering, which could be contributing to the accumulation or reduction in both CSF and PET amyloid levels.

These discordances have been described in previous articles^101–103^, suggesting that they might be caused due to the differential sensitivity to amyloid species across the AD continuum. Further research is needed to elucidate the role of these common and discordant amyloid mechanisms occurring in brain and their impact on disease development.

In this study, we found a limited overlap between genetic modulators of amyloid burden and the proteins associated with the CSF levels of Aβ42. This could be interpreted as a result of the inherent statistical noise in these multiomic analyses, the lack of power in our main analysis, or it could indicate that the observed discordance is genuine. The poor heritability reported for CSF traits in previous studies^9^ supports that common SNPs might not strongly modulate the CSF amyloid burden. Moreover, no amyloid PRS showed a significant association with the risk of developing AD, whereas the AD PRS showed a strong association with the AD case-control status and amyloid levels, which is fully consistent with previous studies^70,104–106^. These results suggest either a lack of statistical power to detect genuine hits associated with amyloid burden or a limited causal role of common genetic modulators of amyloid deposits in the aetiology of clinical AD. Further studies are needed to clarify these discrepancies. Interestingly, we observed a higher number of *loci*/proteins overlapped with the SOMAscan protein associations with CSF Aβ42 levels and the gene-based analysis than in the sentinel SNP-based GWAS analyses (our meta-analysis n=21). The gene-based approach could be particularly powerful because the genetic markers summarised at (protein-coding) gene level might reduce the statistical noise on a full GWAS dataset^52^. We also noted a large number of significant CSF SOMAscan proteins associated with CSF Aβ42 levels. Notably, most of the observed associations were predominantly positive in our study. Interestingly, Bader *et al*^107^ reported a correlation map illustrating high correlations between CSF proteomic measures suggesting that these measures might lead to multiple significant associations. The massive abundance of significant proteins might simply reflect a general neurodegenerative signature that occurs as a result of widespread neuronal cell death or reactive gliosis. These changes are likely to be epiphenomenal rather than specific to the AD process. The potential implication of these findings is important for interpreting CSF proteome results. Indeed, only a minority of proteomic markers associated to Aβ42 might be genuine mediators modulating the AD-related amyloid endophenotype. Overall, the lack of overlap between Aβ42 and AD risk GWAS studies suggests that genetic factors modulating amyloid production may represent only a relatively small component of overall AD causality. These findings are also in line with several clinical trials targeting amyloid, that have observed a reduced association between Aβ reduction and AD progression, as well as only modest control of AD progression with these monotherapies. This also suggests that both amyloid-dependent and amyloid-independent mechanisms must be addressed simultaneously to effectively control disease progression^108,109^.

Despite the poor overlap, we detected 35 overlapping genes and proteins pointing to a few enriched mechanisms in our CSF-PET meta-GWAS. We consider these overlapping signals of special importance because they could point to genuine amyloid-related mechanisms involved in AD causality and development. We found Aβ burden significantly associated with pathways controlling the anchored proteins in the membrane, which had also been represented in the PAD enriched analysis (n=23 loci/proteins). Interestingly, six enriched mechanisms were represented in both overlapping loci/protein rankings of the PAD and our CSF-PET meta-analysis. These results validate our findings and suggest that the enrichment analysis is more powerful in detecting genuine associations than analysing individual genes, particularly in the context of reduced statistical power.

Additionally, the enrichment analysis pointed to synapse molecules and cell adhesion mechanisms. Neuronal cadherins and integrins have been linked to the synaptic process, plasticity and long-term potentiation and modulation of Aβ levels^110^, while their loss has been correlated to cognitive decline^111–113^. Furthermore, we detected a link between amyloid levels and mental disorders, such as anxiety which has been associated with high Aβ deposition across the AD continuum^114–116^. On the contrary, autism spectrum disorder (ASD) has been associated with Aβ processing via the non-amyloidogenic pathway leading to reduced Aβ levels in ASD patients^117^. Other overlapping *loci* and proteins such as *ROBO2*, *CNTN5*, *OPCML*, *NRG3, NGFR* or *CACNA2D3*, have been associated with cognitive performance^118,119^, age at onset^120,121^, schizophrenia^122–124^ or ASD^125,126^, AD^127,128^ and its endophenotypes ^129,130^.

Considering all these observations, it is difficult to conceive that all of them can be explained by pure random chance. However, our analysis had important limitations. First, we use a suboptimal p-value-based meta-analysis method, however, this strategy becomes highly valuable for integrating diverse studies reporting different estimate metrics and combining endophenotypes measured by various techniques^131,132^. Also, the CSF-PET meta-analysis did not report effect size which were estimated. The restrictive SNP filtering allowed the evaluation of only 4.9% of genomic markers, likely due to meta-analysing multiple datasets and reducing marker identification involved in common mechanisms between soluble-CSF and insoluble-PET amyloid species. Moreover, as mentioned earlier, the PAD analysis was not completely independent, with an 8.272% and 3.073% of overlapping samples between our main meta-analysis, the CSF^9^ and PET^48^ summary statistics, respectively. The PAD CSF-PET meta-analysis should be interpreted with extreme caution due to these overlapping samples among summary statistics. Because we used publicly available results, we could not confirm the presence of additional overlapping samples, potentially leading to overfitting. The Ali *et al* meta-GWAS conducted a different data harmonisation process, potentially introducing variability. Furthermore, neuropathological information was not available for these samples, leaving us unaware of other concomitant pathological changes. Finally, the lack of significant findings for several PRS associations may suggest that there is insufficient statistical power to find genetic variants that affect the amyloid endophenotype. These concerns should be addressed in future research.

In summary, our results demonstrate the feasibility of combining Aβ endophenotypes in CSF and PET, along with proteome analysis, to gain novel insights into the fundamental biology of AD. The strong proteomic associations with Aβ endophenotypes could help identify signalling pathways and molecular mechanisms involved in Aβ and AD pathology, as well as the overlapping pathways that control the amyloidotic process. Further studies are needed to refine these observed associations, connecting AD *loci* and proposed causal pathways with brain amyloidogenesis.

## Declarations

### Ethics approval and consent to participate

In accordance with Spanish regulations for the biomedical research field, all the protocols of this study were approved by the Clinical Research Ethics Commission of the Hospital Clinic (Barcelona, Spain) for Ace cohort and the Clinical Research Ethics Commission of Cantabria (Spain) for Valdecilla cohort. This research followed the Declaration of Helsinki. All participants were informed about the procedures and objectives of this study by a neurologist before signing an informed consent. Moreover, data confidentiality and privacy of patients were protected as specified in applicable laws.

## Data Availability

The data that support the findings of this study are publicly available from the corresponding authors upon reasonable request. Additionally, the raw SOMAscan proteomic data is publicly accessible through the Alzheimer's Disease Data Initiative (ADDI) community.

## Acknowledgments

We would like to thank patients and controls who participated in this project. The present work has been performed as part of the doctoral thesis of RPF at the University of Barcelona (Barcelona, Spain). Some control samples and data from patients included in this study were provided in part by the National DNA Bank Carlos III (www.bancoadn.org, University of Salamanca, Spain) and Hospital Universitario Virgen de Valme (Sevilla, Spain); they were processed following standard operating procedures with the appropriate approval of the Ethical and Scientific Committee. Data used in this article were obtained from the Alzheimer’s Disease Neuroimaging Initiative (ADNI) database (adni.loni.usc.edu). As such, the investigators within the ADNI contributed to the design and implementation of ADNI and/or provided data but did not participate in analysis or writing of this report. A complete listing of ADNI investigators can be found at: http://adni.loni.usc.edu/wp-content/uploads/how_to_apply/ADNI_Acknowledgement_List.pdf. Data was used for this project of which collection and sharing was funded by the Alzheimer’s Disease Neuroimaging Initiative (ADNI) (National Institutes of Health Grant U01 AG024904) and DOD ADNI (Department of Defense award number W81XWH-12–2-0012). ADNI is funded by the National Institute on Aging, the National Institute of Biomedical Imaging and Bioengineering, and through generous contributions from the following: AbbVie, Alzheimer’s Association; Alzheimer’s Drug Discovery Foundation; Araclon Biotech; BioClinica, Inc.; Biogen; Bristol-Myers Squibb Company; CereSpir, Inc.; Cogstate; Eisai Inc.; Elan Pharmaceuticals, Inc.; Eli Lilly and Company; EuroImmun; F. Hofmann-La Roche Ltd and its afliated company Genentech, Inc.; Fujirebio; GE Healthcare; IXICO Ltd.; Janssen Alzheimer Immunotherapy Research & Development, LLC.; Johnson & Johnson Pharmaceutical Research & Development LLC.; Lumosity; Lundbeck; Merck & Co., Inc.; Meso Scale Diagnostics, LLC.; NeuroRx Research; Neurotrack Technologies; Novartis Pharmaceuticals Corporation; Pfzer Inc.; Piramal Imaging; Servier; Takeda Pharmaceutical Company; and Transition Therapeutics. The Canadian Institutes of Health Research is providing funds to support ADNI clinical sites in Canada. Private sector contributions are facilitated by the Foundation for the National Institutes of Health (www.fnih.org). The grantee organization is the Northern California Institute for Research and Education, and the study is coordinated by the Alzheimer’s Therapeutic Research Institute at the University of Southern California. ADNI data are disseminated by the Laboratory for Neuro Imaging at the University of Southern California.

## The GR@ACE study group

Nuria Aguilera^1^, Emilio Alarcon^1^, Montserrat Alegret^1,3^, Mercè Boada^1,3^, Mar Buendia^1^, Amanda Cano^1,3^, Pilar Cañabate^1,3^, Angel Carracedo^5,33^, Arturo Corbatón-Anchuelo^23^, Itziar de Rojas^1,3^, Susana Diego^1^, Ana Espinosa^1^, Anna Gailhajenet^1^, Pablo García-González^1,3^, Marina Guitart^1^, Antonio González-Pérez^32^, Marta Ibarria^1^, Asunción Lafuente^1^, Juan Macias^22^, Olalla Maroñas^5^, Elvira Martín^1^, Maria Teresa Martínez Larrad^23,26^, Marta Marquié^1,3^, Laura Montrreal^1^, Sonia Moreno-Grau^1,3^, Mariona Moreno^1^, Raúl Nuñez-Llaves1^1^, Clàudia Olivé^1^, Adelina Orellana^1^, Gemma Ortega^1,3^, Ana Pancho^1^, Ester Pelejà^1^, Alba Pérez-Cordon^1^, Juan A Pineda^16^, Raquel Puerta^1,2^, Silvia Preckler^1^, Inés Quintela^5^, Luis Miguel Real^22,16^, Maitee Rosende-Roca^1^, Agustín Ruiz^1,3^, Maria Eugenia Sáez^32^, Angela Sanabria^1,3^, Manuel Serrano-Rios^23^, Oscar Sotolongo-Grau^1^, Luís Tárraga^1,3^, Sergi Valero^1,3^, Liliana Vargas^1^

^1^Research Center and Memory Clinic. Ace Alzheimer Center Barcelona – Universitat Internacional de Catalunya, Spain. ^2^Universitat de Barcelona (UB). ^3^CIBERNED, Network Center for Biomedical Research in Neurodegenerative Diseases, National Institute of Health Carlos III, Madrid, Spain. ^5^Grupo de Medicina Xenómica, Centro Nacional de Genotipado (CEGEN-PRB3-ISCIII). Universidade de Santiago de Compostela, Santiago de Compostela, Spain. ^16^Departamento de Especialidades Quirúrgicas, Bioquímica e Inmunología. School of Medicine. University of Malaga. Málaga, Spain. ^22^Unidad Clínica de Enfermedades Infecciosas y Microbiología.Hospital Universitario de Valme, Sevilla, Spain. ^23^Instituto de Investigación Sanitaria del Hospital Clínico San Carlos (IdISSC), Hospital Clínico San Carlos. ^26^Centro de Investigación Biomédica en Red de Diabetes y Enfermedades Metabólicas Asociadas (CIBERDEM). ^32^CAEBI, Centro Andaluz de Estudios Bioinformáticos, Sevilla, Spain. ^33^Fundación Pública Galega de Medicina Xenómica – CIBERER-IDIS, Santiago de Compostela, Spain.

## DEGESCO consortium

Astrid Daniela Adarmes-Gómez^28,3^, Miquel Aguilar^36,37^, Nuria Aguilera^1^, Emilio Alarcón-Martín^1^, Daniel Alcolea^31,3^, Montserrat Alegret^1,3^, María Dolores Alonso^38^, Ignacio Alvarez^36,37^, Victoria Álvarez^20,21^, Guillermo Amer-Ferrer^39^, Martirio Antequera^40^, Anna Antonell^30^, Carmen Antúnez^41^, Alfonso Pastor Arias^10,11^, Miquel Baquero^42^, Olivia Belbin^31,3^, María Bernal Sánchez-Arjona^27^, Mercè Boada^1,3^, Mar Buendia^1^, Dolores Buiza-Rueda^28,3^, María Jesús Bullido^17,3,18,19^, Mariateresa Buongiorno^36,37^, Juan Andrés Burguera^42^, Miguel Calero^29,3,43^, Amanda Cano^1,3^, Pilar Cañabate^1,3^, Fernando Cardona Serrate^8,9,3^, Ángel Carracedo^5,33^, Fátima Carrillo^28,3^, María José Casajeros^44^, Jordi Clarimon^31,3^, Arturo Corbatón-Anchuelo^23^, Anaïs Corma-Gómez^22^, Paz de la Guía^15^, Itziar de Rojas^1,3^, Teodoro del Ser^45^, Susana Diego^1^, Mónica Diez-Fairen^36,37^, Oriol Dols-Icardo^31,3^, Ana Espinosa^1^, Marta Fernández-Fuertes^22^, Juan Fortea^31,3^, Emilio Franco-Macías^27^, Ana Frank-García^19,3,46,47^, Anna Gailhajenet^1^, Jose María García-Alberca^15^, Pablo García-González^1,3^, Sebastián García-Madrona^44^, Guillermo Garcia-Ribas^44^, Lorena Garrote-Espina^28,3^, Pilar Gómez-Garre^28,3^, Antonio González-Pérez^32^, Marina Guitart^1^, Raquel Huerto Vilas^10,11^, Marta Ibarria^1^, Silvia Jesús^28,3^, Miguel Angel Labrador Espinosa^28,3^, Asunción Lafuente^1^, Carmen Lage^4,3^, Agustina Legaz^40^, Alberto Lleó^31,3^, Sara López-García^4,3^, Adolfo Lopez de Munain^12,13,3,14^, Juan Macías^22^, Daniel Macias-García^28,3^, Salvadora Manzanares^40^, Marta Marín^27^, Juan Marín-Muñoz^40^, Olalla Maroñas^5^, Marta Marquié^1,3^, Elvira Martín^1^, Angel Martín Montés^47,48^, Begoña Martínez^40^, Victoriana Martínez^40^, Pablo Martínez-Lage Álvarez^49^, María Teresa Martínez-Larrad^23,26^, Marian Martinez de Pancorbo^50^, Carmen Martínez Rodríguez^51,21^, Miguel Medina^3,29^, Maite Mendioroz Iriarte^52^, Silvia Mendoza^15^, Manuel Menéndez-González^21,53^, Pablo Mir^28,3^, Laura Molina-Porcel^30,54^, Laura Montrreal^1^, Mariona Moreno^1^, Fermin Moreno^12,14,3^, Laura Muñoz-Delgado^28,3^, Fuensanta Noguera Perea^40^, Raúl Núñez-Llaves^1^, Clàudia Olivé^1^, Gemma Ortega^1^, Ana Pancho^1^, Ana Belén Pastor^29,55^, Pau Pastor^6,7^, Ester Pelejá^1^, Alba Pérez-Cordón^1^, Jordi Pérez-Tur^8,3,9^, María Teresa Periñán^28,3^, Juan Antonio Pineda^22^, Rocío Pineda-Sánchez^28,3^, Gerard Piñol-Ripoll^10,11^, Silvia Preckler^1^, Raquel Puerta^1,2^, Inés Quintela^5^, Alberto Rábano^29,55,3^, Luis Miguel Real^22^, Diego Real de Asúa^56^, Eloy Rodriguez-Rodriguez^4,3^, Irene Rosas Allende^20,21^, Maitée Rosende-Roca^1,3^, Jose Luís Royo^16^, Agustín Ruiz^1,3^, María Eugenia Sáez^32^, Ángela Sanabria^1,3^, Pascual Sánchez-Juan^4,3^, Raquel Sánchez-Valle^30^, Isabel Sastre^17,3^, Manuel Serrano-Ríos^23^, Oscar Sotolongo-Grau^1^, Lluís Tárraga^1,3^, Sergi Valero^1,3^, Liliana Vargas^1^, María Pilar Vicente^40^, Laura Vivancos-Moreau^40^, Miren Zulaica^14,3^

^1^Research Center and Memory Clinic. Ace Alzheimer Center Barcelona – Universitat Internacional de Catalunya, Spain. ^2^Universitat de Barcelona (UB). ^3^CIBERNED, Network Center for Biomedical Research in Neurodegenerative Diseases, National Institute of Health Carlos III, Madrid, Spain. ^4^Neurology Service, Marqués de Valdecilla University Hospital (University of Cantabria and IDIVAL), Santander, Spain. ^5^Grupo de Medicina Xenómica, Centro Nacional de Genotipado (CEGEN-PRB3-ISCIII). Universidade de Santiago de Compostela, Santiago de Compostela, Spain. ^6^Unit of Neurodegenerative diseases, Department of Neurology, University Hospital Germans Trias i Pujol, Badalona, Barcelona, Spain. ^7^The Germans Trias i Pujol Research Institute (IGTP), Badalona, Barcelona, Spain. ^8^Unitat de Genètica Molecular, Institut de Biomedicina de València-CSIC, Valencia, Spain. ^9^Unidad Mixta de Neurologia Genètica, Instituto de Investigación Sanitaria La Fe, Valencia, Spain. ^10^Unitat Trastorns Cognitius, Hospital Universitari Santa Maria de Lleida, Lleida, Spain. ^11^Institut de Recerca Biomedica de Lleida (IRBLLeida), Lleida, Spain. ^12^Department of Neurology. Hospital Universitario Donostia. San Sebastian, Spain. ^13^Department of Neurosciences. Faculty of Medicine and Nursery. University of the Basque Country, San Sebastián, Spain. ^14^Neurosciences Area. Instituto Biodonostia. San Sebastian, Spain. ^15^Alzheimer Research Center & Memory Clinic, Andalusian Institute for Neuroscience, Málaga, Spain. ^16^Departamento de Especialidades Quirúrgicas, Bioquímica e Inmunología. School of Medicine. University of Malaga. Málaga, Spain. ^17^Centro de Biología Molecular Severo Ochoa (UAM-CSIC). ^18^Instituto de Investigacion Sanitaria ‘Hospital la Paz’ (IdIPaz), Madrid, Spain. ^19^Universidad Autónoma de Madrid. ^20^Laboratorio de Genética. Hospital Universitario Central de Asturias, Oviedo, Spain. ^21^Instituto de Investigación Sanitaria del Principado de Asturias (ISPA). ^22^Unidad Clínica de Enfermedades Infecciosas y Microbiología.Hospital Universitario de Valme, Sevilla, Spain. ^23^Instituto de Investigación Sanitaria del Hospital Clínico San Carlos (IdISSC), Hospital Clínico San Carlos. ^26^Centro de Investigación Biomédica en Red de Diabetes y Enfermedades Metabólicas Asociadas (CIBERDEM). ^27^Dementia Unit, Department of Neurology, Hospital Universitario Virgen del Rocío, Instituto de Biomedicina de Sevilla (IBiS), Sevilla, Spain. ^28^Unidad de Trastornos del Movimiento, Servicio de Neurología y Neurofisiología.Instituto de Biomedicina de Sevilla (IBiS), Hospital Universitario Virgen del Rocío/CSIC/Universidad de Sevilla, Seville, Spain. ^29^CIEN Foundation/Queen Sofia Foundation Alzheimer Center. ^30^Alzheimer’s disease and other cognitive disorders unit.Service of Neurology.Hospital Clínic of Barcelona. Institut d’Investigacions Biomèdiques August Pi i Sunyer, University of Barcelona, Barcelona, Spain. ^31^Department of Neurology, Sant Pau Memory Unit, Sant Pau Biomedical Research Institute, Hospital de la Santa Creu i Sant Pau, Universitat Autònoma de Barcelona,Barcelona, Spain. ^32^CAEBI, Centro Andaluz de Estudios Bioinformáticos, Sevilla, Spain. ^33^Fundación Pública Galega de Medicina Xenómica – CIBERER-IDIS, Santiago de Compostela, Spain. ^36^Fundació Docència i Recerca MútuaTerrassa, Terrassa, Barcelona, Spain. ^37^Memory Disorders Unit, Department of Neurology, Hospital Universitari Mutua de Terrassa, Terrassa, Barcelona, Spain. ^38^Servei de Neurologia.Hospital Clínic Universitari de València. ^39^Department of Neurology, Hospital Universitario Son Espases, Palma, Spain. ^40^Unidad de Demencias.Hospital Clínico Universitario Virgen de la Arrixaca, Palma, Spain. ^41^Unidad de Demencias, Hospital Clínico Universitario Virgen de la Arrixaca, Murcia, Spain. ^42^Servei de Neurologia, Hospital Universitari i Politècnic La Fe, Valencia, Spain. ^43^UFIEC, Instituto de Salud Carlos III. ^44^Hospital Universitario Ramon y Cajal, IRYCIS, Madrid, Spain. ^45^Department of Neurology/CIEN Foundation/Queen Sofia Foundation Alzheimer Center. ^46^Department of Neurology, La Paz University Hospital. Instituto de Investigación Sanitaria del Hospital Universitario La Paz. IdiPAZ. ^47^Hospital La Paz Institute for Health Research, IdiPAZ, Madrid, Spain. ^48^Department of Neurology, La Paz University Hospital. ^49^Centro de Investigación y Terapias Avanzadas. Fundación CITA-Alzheimer, San Sebastián, Spain. ^50^BIOMICS País Vasco; Centro de investigación Lascaray, Universidad del País Vasco UPV/EHU, Vitoria-Gasteiz, Spain. ^51^Hospital de Cabueñes, Gijón, Spain. ^52^Navarrabiomed, Pamplona, Spain. ^53^Departamento de Medicina, Universidad de Oviedo, Oviedo, Spain. ^54^Neurological Tissue Bank of the Biobanc-Hospital Clinic-IDIBAPS, Institut d’Investigacions Biomèdiques August Pi i Sunyer, Barcelona, Spain. ^55^BT-CIEN. ^56^Hospital Universitario La Princesa, Madrid, Spain.

## The FACEHBI study group

JA Alllué^57^, F Appiani^1^, DM Ariton^1^, M Berthier^58^, U Bojaryn^1^, M Buendia^1^, S Bullich^59^, F Campos^60^, P Cañabate^1,3^, L Cañada^1^, C Cuevas^1^, S Diego^1^, JM Escudero^61^, A Espinosa^1,3^, A Gailhajenet^1^, J Giménez^61^, M Gómez-Chiari^61^, M Guitart^1^, I Hernández^1,3^, M Ibarria^1^, A Lafuente^1^, N Lleonart^1^, F Lomeña^60^, E Martín^1^, M Moreno^1^, A Morera^1^, N Muñoz^1^, A Niñerola^60^, AB Nogales^1^, L Núñez^62^, G Ortega^1,3^, A Páez^62^, A Pancho^1^, E Pelejà^1^, E Pérez-Martínez^59^, A Pérez-Cordon^1^, V Pérez-Grijalba^57^, M Pascual-Lucas^57^, A Perissinotti^60^, S Preckler^1^, M Ricciardi^1^, N Roé-Vellvé^59^, J Romero^57^, MI Ramis^1^, M Rosende-Roca^1^, M Sarasa^57^, S Seguer^1^, A Stephens^59^, MA Tejero^61^, J Terencio^57^, M Torres^62^, L Vargas^1^, A Vivas-Larruy^61^

^1^Research Center and Memory Clinic. Ace Alzheimer Center Barcelona – Universitat Internacional de Catalunya, Spain. ^3^CIBERNED, Network Center for Biomedical Research in Neurodegenerative Diseases, National Institute of Health Carlos III, Madrid, Spain. ^57^Araclon Biotech-Grífols.Zaragoza, Spain. ^58^Cognitive Neurology and Aphasia Unit (UNCA). University of Malaga. Málaga, Spain. ^59^Life Molecular Imaging GmbH. Berlin, Germany. ^60^Servei de Medicina Nuclear, Hospital Clínic i Provincial. Barcelona, Spain. ^61^Departament de Diagnòstic per la Imatge. Clínica Corachan,

## Author contributions

ACF, IdR and AR designed and conceptualized the study and interpreted the data. RP, AR and IdR contributed to data acquisition, analysis, interpreted the data and co-wrote the manuscript. PGG, CO, OSG contributed to data interpretation. AR supervised the study. MA, SV, MMS, MB, PSJ, ACS, ACF, JEC, AR contributed to the critical revision of the paper. All authors critically revised the manuscript for important intellectual content and approved the final manuscript. GR@ACE/DEGESCO Data generation: RP, IdR, PGG, CO, AGS, FGG, LM, CL, IQ, NA, ERR, EAM, AO, AC, MES and ACF. Sample contribution: RP, IdR, PGG, CO, AGS, FGG, LM, VP, CL, IQ, NA, ERR, EAM, AO, PP, JPT, GPR, ALM, JMGA, JLR, MJB, VA, LMR, ACA, DGG, MML, EFM, PM, MM, ODI, LT, MA, SV, MMS, MB, PSJ, ACS, ACF and AR. Analysis: RP and IdR. Study supervision/management: LT, MB, PSJ, ACF and AR.

## Abbreviations

AD: Alzheimer’s disease
ADNI: Alzheimer’s Disease Neuroimaging Initiative
ASD: autism spectrum disorder
Aβ: amyloid
Aβ42: amyloid beta 42
AV45: Florbetapir
CADD: Combined Annotation Dependent Depletion
CDR: Clinical Dementia Rating
CI: Confidence interval
eQTL: expression quantitative trait loci
FBB: Florbetaben
FDR: false discovery rate
FUMA: Functional Mapping and Annotation of Genome-Wide Association Studies
GPI: glycosylphosphatidyl inositol
GWAS: Genome-wide association studies
HC: Healthy Control
LP: lumbar puncture
MAC: minor allele count
MAF: minor allele frequency
MCI: Mild Cognitive impairment
MMSE: Mini-Mental State Exam
n: Sample size
NIA-AA: National Institute on Aging and Alzheimer’s Association
NINCDS/ADRDA: National Institute of Neurological and Communicative Disorders and Stroke and Alzheimer’s Disease and Related Disorders Association
OR: Odds ratio
P: p-value
p-tau: phosphorylated tau in Thr 181
PAD: publicly available datasets
PET: Positron Emission Tomography
PCA: principal component analysis
PCs: Principal components
pQTL: protein quantitative trait loci
PRS: Polygenic risk scores
RFU: relative fluorescent units
SCD: subjective cognitive decline
SNP: single nucleotide polymorphisms
t-tau: total tau
λ: genomic inflation factor

