## Supplementary Material and Figures for "Connecting genomic and proteomic signatures of amyloid burden in the brain"

### Supplementary Figures

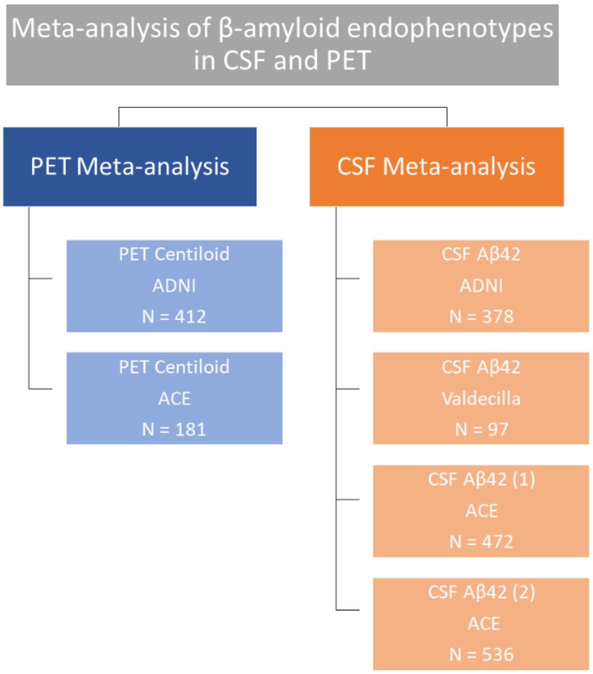


***Supplementary Figure 1. Workflow of this study for each cohort and endophenotypes.*** *ACE cohort used both Innotest ELISA kits (1) and CLEIA Lumipulse (2) for measuring CSF Aβ42 endophenotypes.*

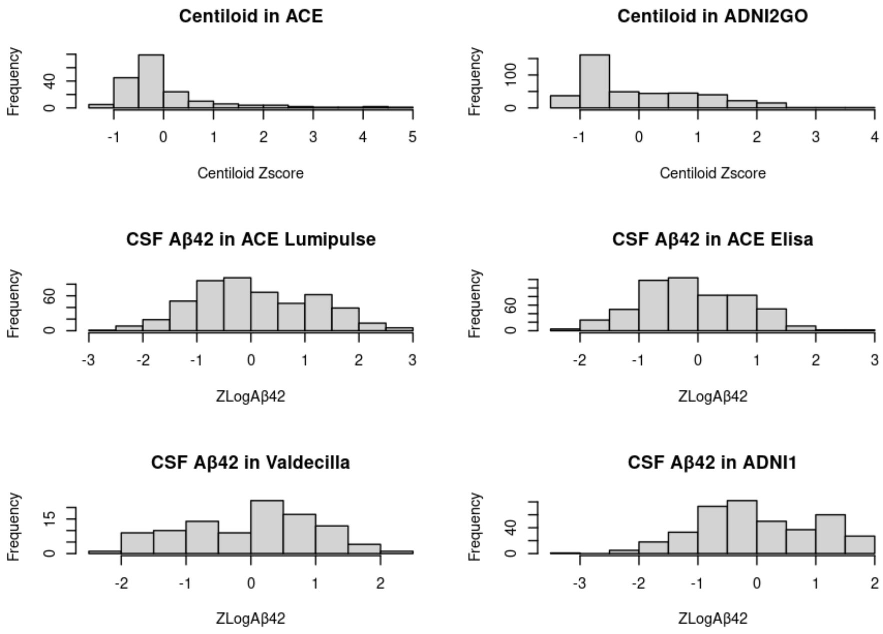


***Supplementary Figure 2. Histograms of the CSF and PET endophenotypes distribution after the data harmonization.***

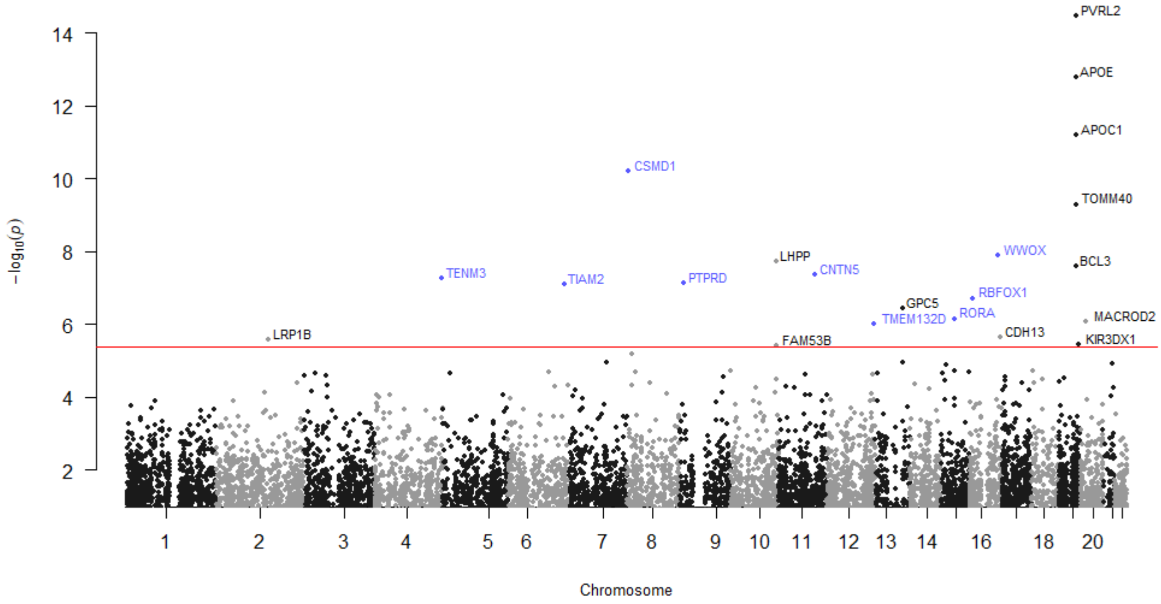


***Supplementary Figure 3. Manhattan plot of the gene-based analysis of MAGMA (FUMA).*** *Genes highlighted in blue were previously related to AD and its endophenotypes. Red line: significance threshold at P<4.235e-06.*

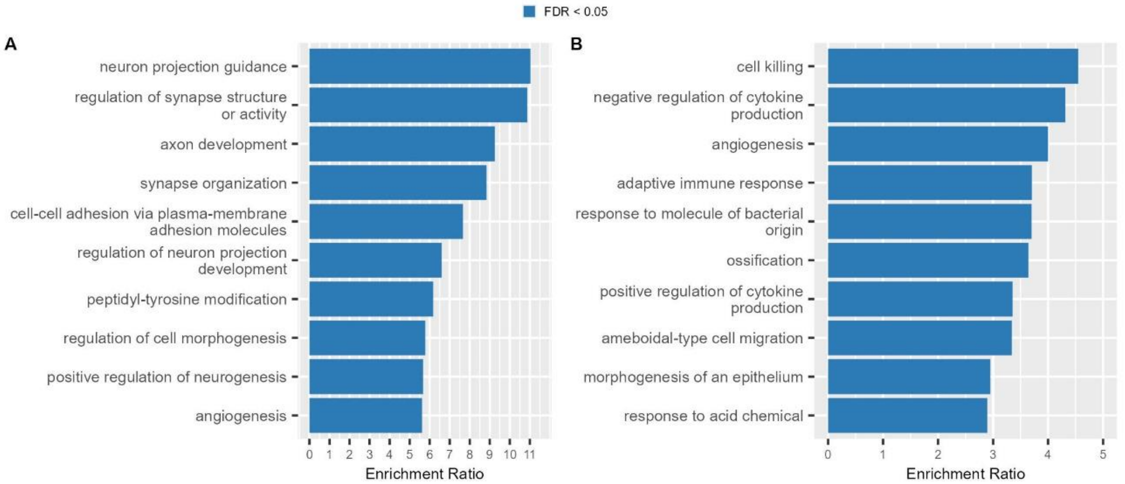


***Supplementary Figure 4. Top10 enriched mechanisms of the ORA analysis using the WebGestalt tool****. A) Considering proteins significantly associated with CSF Aβ42 levels (n=1,387), and B) considering the complete set of proteins (n=2,648) included in CSF proteome analyses.*

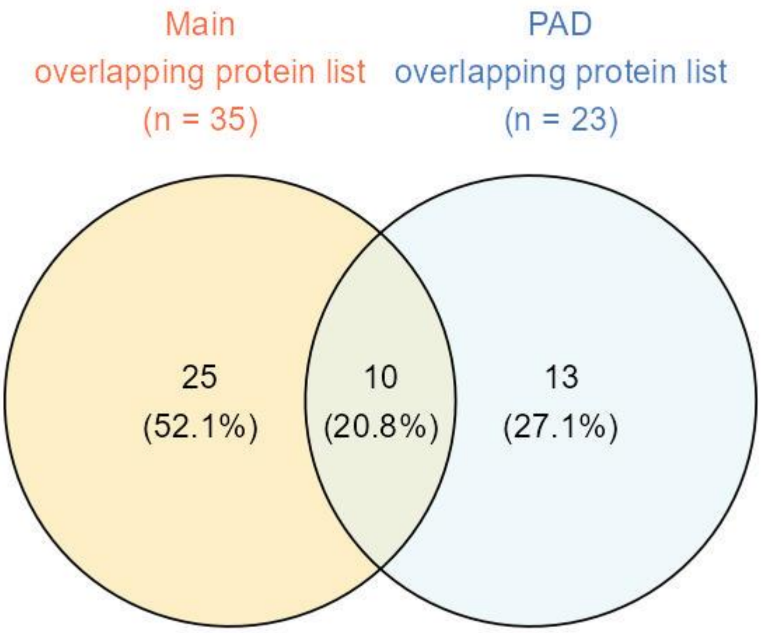


***Supplementary Figure 5.*** *Venn diagram of the overlapping loci/proteins in genomic and proteomic analysis considering the PAD (light blue) and our CSF-PET meta-analysis (yellow).*

### Supplementary Material

#### 1. AD risk and reported amyloidosis genes in the literature

We analysed the effect of AD genes described in Bellenguez *et al* in our Aβ endophenotype study. A total of 12 genes were not considered for replication due to low frequency or missingness in the imputation process. Although the extensive filter of genetic markers did not allow us to evaluate all markers, rs679515-*CR1* was significantly replicated in our amyloid burden meta-analysis ($P$<0.05). Interestingly, this variant was not significant independently in CSF or PET meta-GWAS, but the combination of Aβ endophenotypes allowed us to gain power and replicate it. Additionally, the genetic marker located in the rs6943429-*UMAD1* gene had a consistent effect in the combined Aβ burden meta-analysis but did not reach significance. Moreover, the rs113706587-*RASGEF1C* loci were only significantly replicated in the PET meta-analysis. In contrast, both rs6014724-*CASS4* and rs6605556-*HLA-DQA1* markers were significantly replicated in the CSF Aβ42 meta-analysis and also had a consistent effect size direction in the PET centiloid meta-analysis. Thirty-five and thirty-four additional genetic markers had a consistent effect with what was observed in Bellenguez *et al* study in CSF Aβ42 and PET centiloid meta-analysis, respectively (Supplementary Table 2).

Regarding the genetic variants reported in Jansen *et al* for amyloidosis, the rs429358-*APOE* variant was significantly replicated in all Aβ meta-analyses while the rs4844610-*CR1* variant had a consistent effect in all meta-analyses: it was nominally replicated in CSF Aβ42 and significantly replicated in the final Aβ burden combined meta-analysis (P<0.05; Supplementary Table 3). Finally, we also aimed to replicate markers associated to neuropathological features described in Beecham *et al*. The rs6857 and rs429358 were significantly replicated in all meta-analysis as well as the meta-analysis of CSF-PET endophenotypes. These variants were both located in the *APOE* region and associated with NFTs, neuritic plaques and other phenotypes (Supplementary Table 4).
